# Dynamics of SARS-CoV-2 seroassay sensitivity: a systematic review and modeling study

**DOI:** 10.1101/2022.09.08.22279731

**Authors:** Nana Owusu-Boaitey, Timothy W. Russell, Gideon Meyerowitz-Katz, Andrew T. Levin, Daniel Herrera-Esposito

## Abstract

**Background:** Serological surveys have been the gold standard to estimate the numbers of SARS-CoV-2 infections, epidemic dynamics, and disease severity. Serological assays have decaying sensitivity with time that can bias their results, but there is a lack of guidelines to account for this phenomenon for SARS-CoV-2.

**Aim:** Our goal is to assess the sensitivity decay of seroassays for detecting SARS-CoV-2 infections, the dependence of this decay on assay characteristics, and to provide a simple method to correct for this phenomenon.

**Methods:** We performed a systematic review and meta-analysis of SARS-CoV-2 serology studies. We included studies testing previously diagnosed individuals, without any SARS-CoV-2 vaccines, and excluded studies of cohorts highly unrepresentative of the general population (e.g. hospitalised patients).

**Results:** Of the 488 screened studies, 76 studies reporting on 50 different seroassays were included in the analysis. Sensitivity decay depends strongly on the antigen and the analytic technique used by the assay, with average sensitivities ranging between 26% and 98% at 6 months after infection, depending on assay characteristics. We find that a third of the included assays depart considerably from manufacturer specifications after 6 months.

**Conclusions:** Seroassay sensitivity decay depends on assay characteristics, and for some types of assays it can make manufacturer specifications highly unreliable. We provide a tool to correct for this phenomenon, and to assess the risk of decay for a given assay. Our analysis can guide the design and interpretation of serosurveys for SARS-CoV-2 and other pathogens, and quantify systematic biases in the existing serology literature.

## Introduction

Throughout the SARS-CoV-2 pandemic, policymakers have been guided by the inferred number of past infections based on serological assays. Seroassays have been heavily used to estimate the proportion of individuals that have been infected, the rate of fatal or severe infections (1–5); population-wide immunity (6–8); and to anticipate the effect of future infection waves (9,10), among others.

However, antibody levels wane with time after infection (11), reducing the sensitivity of serological assays for detecting previous infections (12–14). We refer to the decay of assay sensitivity (in the context of serosurveillance) with time after seroconversion as seroreversion (by ‘time’, we refer to the time spanned between COVID-19 diagnosis and serological testing). Seroreversion is a major potential source of bias when estimating numbers of infections (1,15,16), and because these estimates guide public health policies such as vaccination programs, it is important to account for this phenomenon.

More broadly, understanding seroreversion in general is important for the management of other emerging infectious diseases. For this, the study SARS-CoV-2 presents a unique opportunity. First, an emergent pathogen with distinct symptoms, widespread diagnosis, and short incubation times allows for precise timing of epidemic waves and infections. Second, in some cohorts it can be assumed that reinfections are rare (i.e. serosurveys performed after first epidemic waves). Third, large numbers of serological surveys were performed for SARS-CoV-2, using a wide range of assays and cohorts. These features of the SARS-CoV-2 pandemic allow for a rich analysis of seroreversion.

Strikingly, there is a lack of general analyses and guidelines to correct for seroreversion in the SARS-CoV-2 literature, to the best of our knowledge (15,16,19). Time-varying sensitivity of seroassays has been evaluated in previous studies, but these are limited to few assays or short time spans (13,14,20–23). Other studies analysed the change in quantitative antibody levels (11,13,24–28), which is informative for other uses of serological assays (e.g. studying immune protection), but not for infection surveys.

Here we perform a systematic review and meta-analysis of serology studies of COVID-19, to better characterise seroreversion across assays. We collected and curated time-specific sensitivity estimates from serological studies testing previously diagnosed COVID-19 patients who had not received COVID-19 vaccines. We analysed 76 of over 400 screened studies, encompassing 50 seroassays, 290 data points, and 44992 tests.

We present time-varying sensitivity estimates for the assays included in the analysis, and also the dependence of seroreversion on assay characteristics. Our results can also be used to approximate the seroreversion of seroassays not included in our sample. Finally, we compare time-varying sensitivities to manufacturer reported sensitivities, and estimate the risk of seroreversion bias in the literature, providing an overview of how seroreversion impacted the performance of emergency-approved seroassays during the SARS-CoV-2 pandemic.

## Methods

### Literature search

We performed a systematic literature review of seroprevalence studies including studies identified up to July 13, 2022 using search parameters detailed in a prior publication (29).

We supplemented this analysis with a search on Medrxiv, Biorxiv, PubMed, SSRN, and Google Scholar, on June 30, 2021 using the key “COVID-19 longitudinal, antibody waning” and on February 15, 2022 using the key “COVID-19 seroreversion”. Additional studies were taken from a prior review (30). If a study cited prior publications assessing seroreversion in the same research cohort, we included those prior publications.

Inclusion and exclusion criteria for studies to be included in the analysis are listed in **Supplementary A**. The results of the systematic search are summarised in **Figure 1**. Broadly, we excluded studies reporting on vaccinated individuals, on highly unrepresentative groups. Details of the included study cohorts (e.g. age, sex) are shown in **Table S2** and further discussed in **Supplementary A**. Most cohorts (90%) were serologically tested during 2020, indicating that reinfection incidence is likely low in the analysed data (31), and that infections mainly correspond to the original SARS-CoV-2 variant (32). A list of the included studies and search details is presented in the GitHub repository associated with the project.

**Figure 1.**
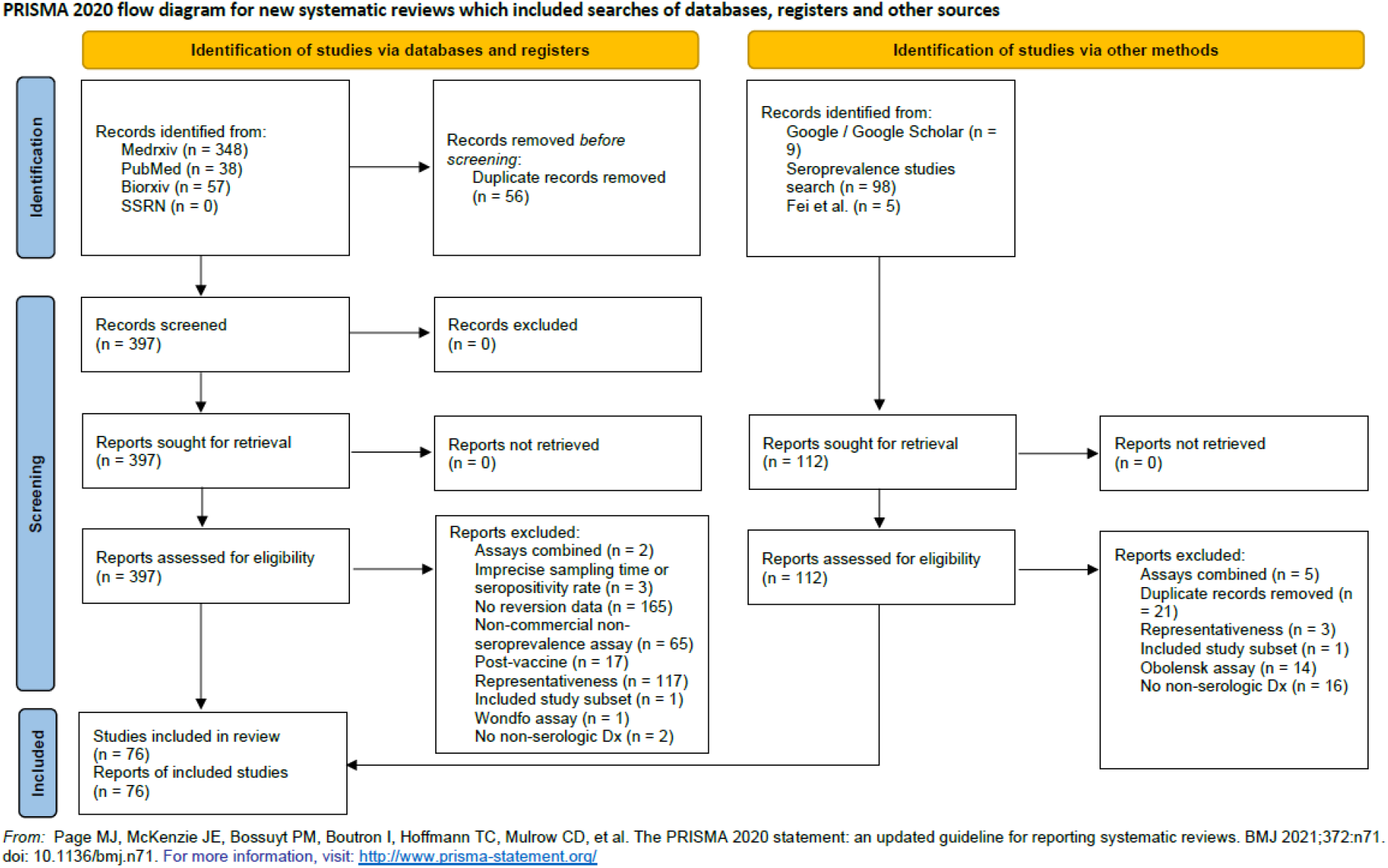
Prisma flow diagram. Flow diagram of the considered studies, showing excluded and included studies.

### Assay characteristics analysed

Serological assays have different characteristics. We considered only some assay characteristics to keep model complexity low. We did not consider antibody isotype because IgG is used in all assays, and preliminary did not show effects for including other isotypes (data not shown). We considered whether the assay is quantitative or a lateral flow assay (LFA). We did not consider the specific type of quantitative readout technique, guided by preliminary analyses (data not shown). We considered all 3 antigens: nucleocapsid, spike protein, and S1-receptor binding protein (RBD). We considered 3 different types of antibody binding in quantitative assays: indirect, competitive and direct (the latter also called double-antigen sandwich assays in the literature).

### Statistical model

We fitted a hierarchical logistic regression Bayesian model to the data. For a given cohort of *N* serologically tested individuals in a study (all which have a previous COVID-19 diagnosis), we model the likelihood of the number of positive results *x*, with a binomial distribution with sensitivity *θ*:

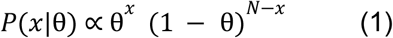

Each cohort of *N* individuals tested in a study is associated with a time of testing *t* (i.e. the average time spanned between COVID-19 diagnosis and serological testing for this cohort). Throughout the text, we refer to a cohort of individuals tested in a given study *s*, at a given time *t*, with an assay *a*, as a data point (e.g. a cohort tested across different times corresponds to multiple data points). We model the sensitivity of data point *θ*_*a,s,t*_ (assay *a*, time *t*, study *s*) with the logit function:

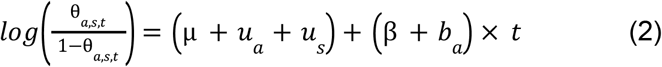

Where *μ* is the mean intercept, *u*_*a*_ and *u*_*s*_ are the random effects on the intercept of assay and study, *β* is the mean time-slope, and *b*_*a*_ is the random effect of assay on the slope. We set flat priors for *μ* and *β*. We set gamma priors with shape and rate parameters of 4 for the standard deviations *σ*_*ua*_, *σ*_*us*_ and *σ*_*ba*_ of the random effects.

To study the effect of assay characteristics, we modify equation (2) to include their effects on the slope:

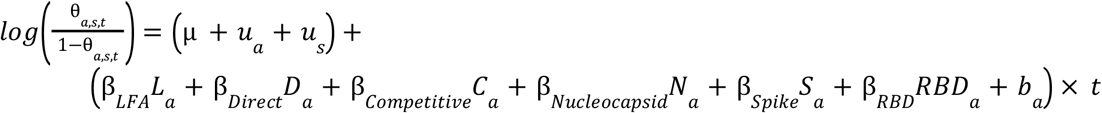

Parameters *β*_*LFA*_, *β*_*Direct*_ and *β*_*Competitive*_ are the effects on the time slope of using, respectively, LFA, Quantitative-Direct, or Quantitative-Competitive assay designs. Variables *L*_*a*_, *D*_*a*_ and *C*_*a*_ take values of 0 or 1 to indicate whether assay *a* uses that design. We do not include an effect for the Quantitative-Indirect design, making it the baseline slope (thus, the parameters above indicate a difference relative to this design). Similarly, *β*_*Nucleocapsid*_, *β*_*Spike*_ and *β*_*RBD*_ are the effects of the antigen used on the time slope.

We fitted the models using STAN (33), with 4 chains with 4000 draws each (1000 warmup) and default parameters.

We tested the model fits using a cross-validation analysis, leaving out data points from model fitting and obtaining sensitivity predictions for the left-out data. We repeated this procedure to obtain an estimate for every data point. We used a tailored procedure that required that every prediction involved extrapolation of the model through time (details in **Supplementary B**).

### Estimation of testing times

When studies did not report the median time between diagnosis and serological testing for their cohort, we estimated these times using reported case curves for the study’s location (see details in **Supplementary A**).

### Data and code availability

All the data, code, literature pointers and review comments are available at the associated GitHub page https://github.com/dherrera1911/seroreversion_metaanalysis.

## Results

### Assay variability in seroreversion

First, we fitted a model without considering assay characteristics. In **Figure 2** we see the slope of sensitivity decay obtained for each assay (the corresponding sensitivity-time curves are shown in **Figure S1**). Estimated slopes were highly variable across assays (random effects of the assay were *σ*_*ua*_=0.26 (95%CrI: 0.19; 0.36) for the intercept and *σ*_*ba*_=0.66 (95%CrI: 0.31; 1.04) for the slope). Interestingly, although most assays had decreasing sensitivity as expected (negative slopes), some assays had increasing sensitivities (positive slopes, shaded region in **Figure 2**). The positive slopes are not due to a lower starting sensitivity, or an initial increase followed by a decay (**Figure S2** shows that both early and late changes in sensitivity are increasing). Because reinfection incidence is likely low in our data, it is unlikely that these results reflect infections. There was also considerable variability in the intercepts between different studies using a same assay, with a standard deviation of *σ*_*us*_=0.81 (95%CrI: 0.67; 0.97) (larger than the between-assay standard deviation), outlining the importance of this source of variability.

**Figure 2.**
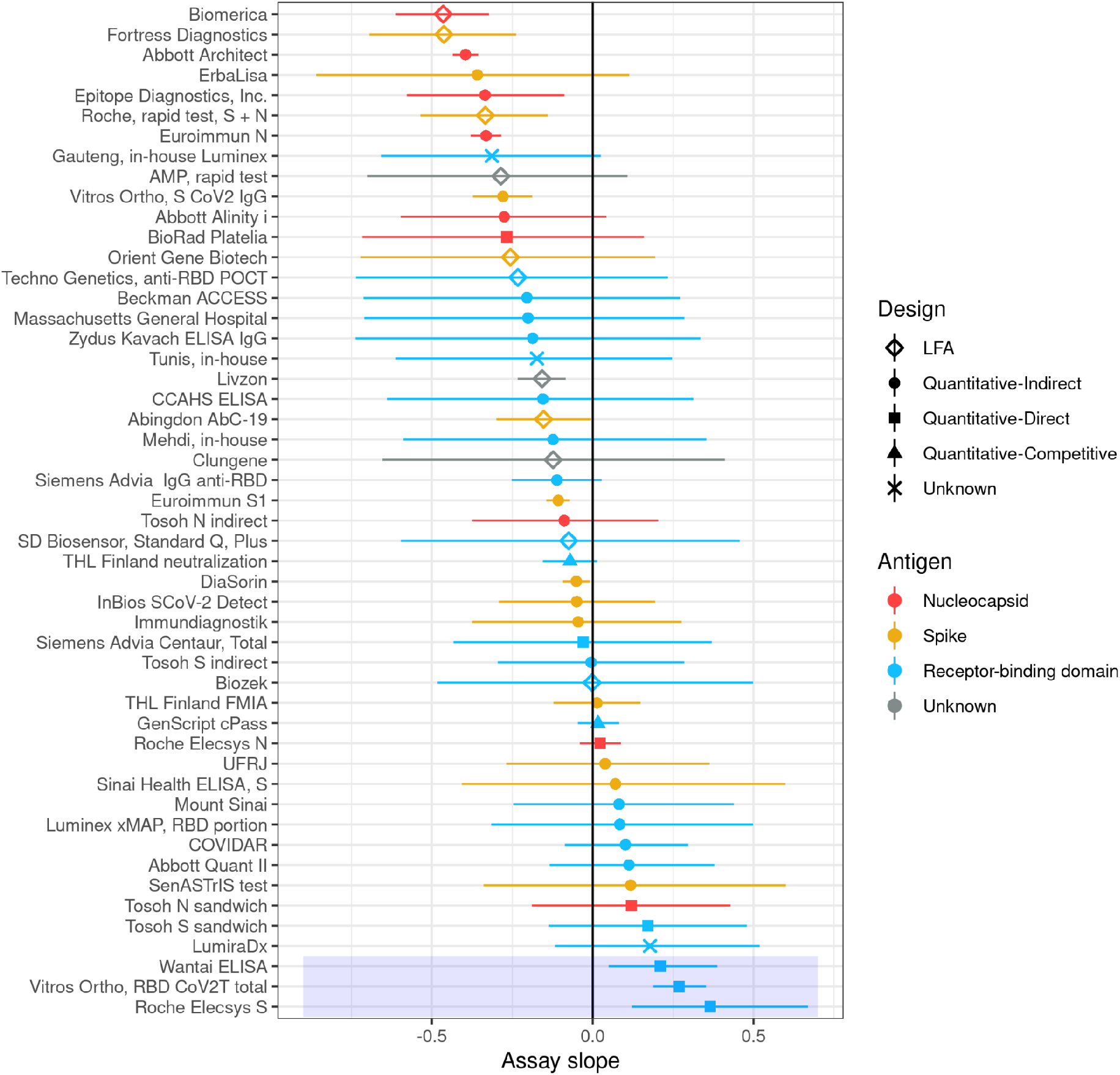
Assay slopes estimated without assay characteristics. Points show the mean of the posterior distribution of the slope for each assay, and horizontal bars show the 95% CrI. The slopes were obtained from the model that does not include effects of assay characteristics, but colour and point shape indicate the characteristics of the assays. The blue rectangle marks assays with slopes significantly greater than 0.

We note that while some assays had many data points spanning several months, other assays only had a few time points (several assays with only a few data points can be seen in **Figure S1**). For the latter, our models sensitivity estimates involve extrapolation of sensitivity across time. We tested our model’s performance at extrapolation using a cross-validation procedure specifically designed for this (method details in **Supplementary B**). We found that the 95% credible intervals (CrI) contained the validation data 91.7% of the time. For assays with fewer than 9 data points (which comprise 99 of the 286 data points), 95.1% of the data points were within the cross validation CrI.

### Assay characteristics determine seroreversion

Next, we analysed the relation between assay characteristics and sensitivity decay. We fitted a model with effects of different assay characteristics on the assay-specific slope. We included terms for each of the 3 antigens (nucleocapsid, spike, and RBD), and for 3 different assay designs (LFA, Quantitative-Direct, Quantitative-Competitive), leaving the fourth assay design (Quantitative-Indirect) as the baseline slope.

Both the analytic technique and the antigen show important effects on seroreversion (**Figure 3, Table 1**). The slope term for LFA assays was negative *β*_*LFA*_=-0.23 (95%CrI: -0.40;-0.07), and *β*_*LFA*_<0 in 99.6% of the posterior samples, indicating that their sensitivity decays faster than Quantitative-Indirect assays. The slope term for Quantitative-Direct assays had a value of *β*_*Direct*_=0.31 (95%CrI: 0.15;0.48), and *β*_*Direct*_>0 in 99.9% of the posterior samples, indicating that they decay more slowly. The term for Quantitative-Competitive assays had a value of *β*_*Competitive*_=-0.03 (95%CrI: -0.25;0.20), and *β*_*Competitive*_>0 in 40.1% of the posterior samples, showing no clear difference with the Quantitative-Indirect assays (which may be due to the small number of assays in the Quantitative-Competitive group). Differences between analytic techniques can be appreciated by comparing the different columns of **Figure 3**.

**Table 1.** Estimated sensitivities (in %) at each time after diagnosis, for each type of assay fitted in the analysis. Each row corresponds to a different type of test, with characteristics indicated in the first 3 columns. Parenthesis show the 95% CrI for each estimate. These estimates do not include the between-assay or between-study variability in their credible intervals. These sensitivities correspond to the red traces and shaded regions in **Figure 3**.

| Month | Assay type |  |  |  |  |  |  |  |  |
| --- | --- | --- | --- | --- | --- | --- | --- | --- | --- |
|  | LFA | LFA | LFA | Q-Indirect | Q-Indirect | Q-Indirect | Q-Direct | Q-Direct | Q-Competitive |
|  | N | S | RBD | N | S | RBD | N | RBD | RBD |
| 1 | 84 (79-89) | 87 (83-91) | 88 (84-91) | 87 (83-91) | 89 (86-92) | 90 (87-93) | 90 (87-93) | 93 (90-95) | 90 (86-93) |
| 2 | 75 (66-83) | 83 (76-88) | 85 (78-91) | 83 (77-88) | 88 (84-92) | 90 (87-93) | 90 (85-94) | 95 (92-96) | 90 (85-93) |
| 3 | 64 (49-76) | 78 (67-86) | 82 (71-90) | 78 (69-85) | 87 (82-92) | 90 (86-94) | 90 (84-94) | 96 (94-98) | 89 (82-94) |
| 4 | 50 (32-69) | 71 (57-83) | 78 (61-89) | 72 (60-82) | 86 (80-91) | 90 (84-94) | 90 (82-95) | 97 (95-98) | 89 (80-95) |
| 5 | 37 (19-60) | 64 (45-80) | 73 (51-89) | 65 (50-78) | 85 (77-91) | 90 (83-95) | 89 (80-95) | 98 (96-99) | 88 (76-95) |
| 6 | 26 (10-50) | 57 (34-77) | 69 (40-89) | 57 (39-74) | 84 (73-91) | 90 (81-95) | 89 (77-96) | 98 (96-99) | 88 (73-96) |
| 7 | 18 (5-41) | 49 (24-73) | 63 (31-88) | 49 (30-70) | 82 (70-92) | 90 (79-96) | 89 (74-97) | 99 (97-100) | 87 (69-97) |
| 8 | 12 (2-32) | 42 (17-69) | 58 (22-88) | 42 (22-65) | 81 (65-92) | 89 (77-96) | 88 (71-97) | 99 (97-100) | 86 (64-97) |
| 9 | 8 (1-24) | 35 (11-65) | 53 (16-88) | 35 (15-60) | 79 (61-92) | 89 (75-97) | 88 (68-97) | 99 (98-100) | 86 (59-97) |
| 10 | 5 (1-18) | 29 (7-61) | 48 (11-87) | 29 (10-55) | 77 (56-92) | 89 (72-97) | 87 (64-98) | 99 (98-100) | 85 (54-98) |
| 11 | 3 (0-13) | 24 (5-57) | 44 (7-87) | 23 (7-50) | 76 (51-92) | 89 (69-98) | 87 (60-98) | 100 (98-100) | 84 (49-98) |
| 12 | 2 (0-9) | 19 (3-53) | 40 (5-86) | 18 (5-45) | 74 (46-92) | 88 (67-98) | 86 (56-98) | 100 (99-100) | 83 (44-98) |
| 13 | 1 (0-6) | 16 (2-48) | 36 (3-86) | 15 (3-40) | 72 (42-92) | 88 (64-98) | 86 (52-99) | 100 (99-100) | 82 (40-99) |

**Figure 3.**
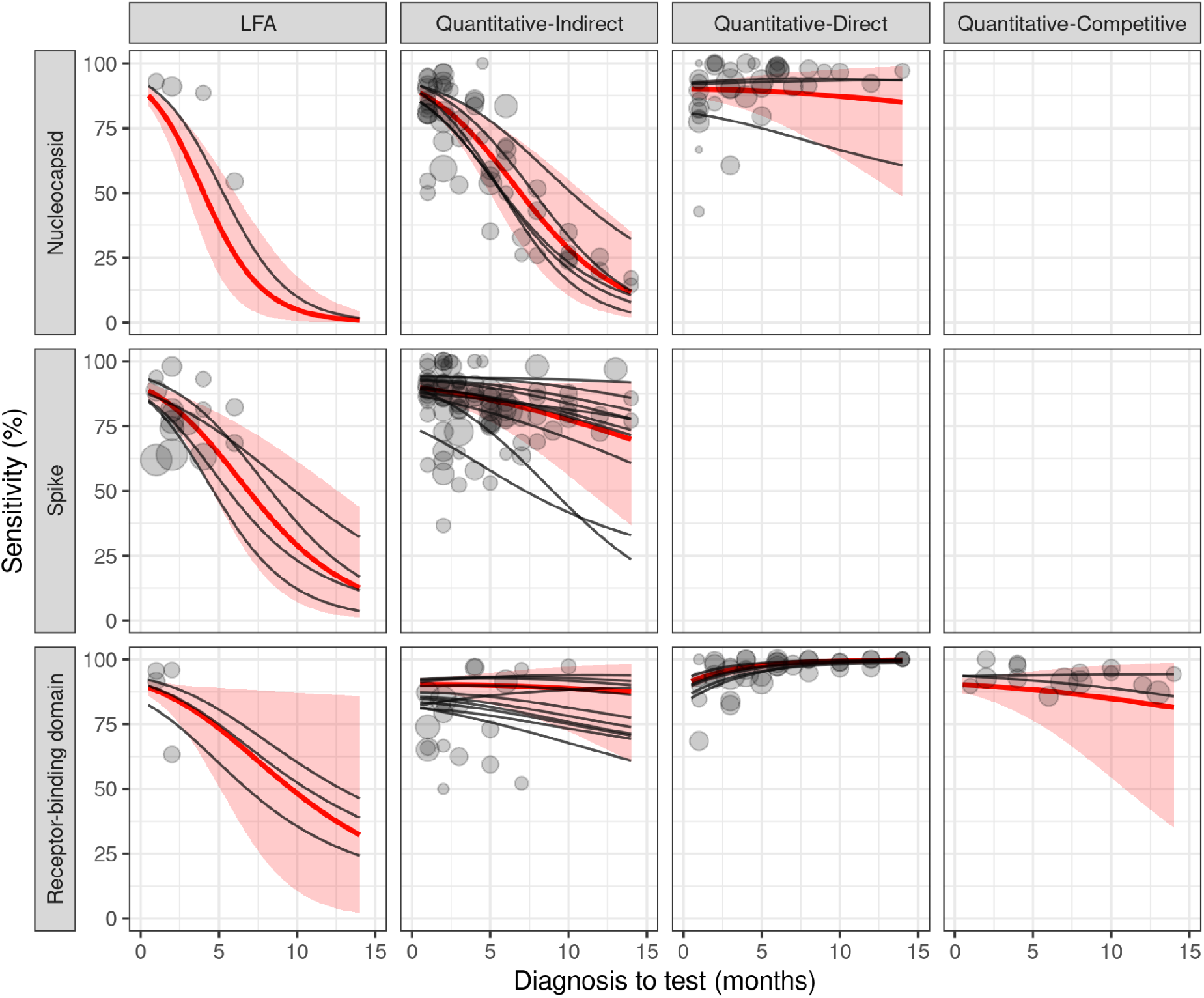
Sensitivity profiles for different assay characteristics. The sensitivity profile across time for each kind of assay is shown in a different panel. Rows indicate the targeted antigen (labelled on the left), and columns indicate the analytic technique (labelled on top). For example, the panel in the second row and second column shows assays that target the spike protein (second row label), and that use a Quantitative-Indirect design (second column label). The red lines show the mean sensitivity for each kind of assay. Shaded regions show the 95% CrI of the mean sensitivity of the group (i.e. not accounting for variability between assays). Black lines show the fits for individual assays. Grey dots show the data, with size proportional to the square root of sample size. Empty panels indicate that no assays with those characteristics were found for the analysis.

On the antigen effect, nucleocapsid targeting assays showed faster seroreversion than spike protein (*β*_*Nucleocapsid*_<*β*_*Spike*_ in 99.7% of the posterior samples). Assays targeting RBD had on average slower seroreversion than spike protein, although the effect was not statistically significant (*β*_*Spike*_<*β*_*RBD*_ in 87.3% of the samples). Differences between antigens can be appreciated by comparing the different rows of **Figure 3**.

To see how the different slopes translate to differences in sensitivity, the reader can compare the sensitivities of the different types of assays for a given month in **Table 1**. Note that there is considerable variability between different assays of the same type (i.e. between the black lines within a same panel). We provide assay-specific sensitivity profiles in **Table S2**. When estimating the extent of seroreversion for a given survey, assay-specific sensitivity estimates should be preferred over the coarser estimates provided for assay types.

Interestingly, we find that one type of assay, the Quantitative-Direct assays targeting RBD-binding antibodies (which contain the 3 assays with statistically significant positive slopes from **Figure 2**) has an average positive slope in 99.9% of the posterior samples. This is in line with previous studies reporting assays of this type to have increasing sensitivity across time, attributing this effect to prolonged antibody maturation (13,14).

All these results were robust to fitting each assay characteristic separately (**Figures S3, S4**), and to excluding data points with estimated times from the model fit (**Figures S5, S6**). The full model had cross-validation accuracy similar to the original model, with data points falling in the 95% CrI of their predictions 92.0% of the time, credible intervals were narrower (more precise) in 81% of the data points.

Finally, we tested whether specificity is also related to assay characteristics. Since specificity does not have temporal dynamics, we only analysed point estimates (see the details of the model in **Supplementary G**). Similar to sensitivity, we found that LFA assays have on average smaller specificities than quantitative assays (*β*_*LFA*_<0 in 98.4% of the posterior samples). Unlike sensitivity, we did not find significant differences with quantitative assays (e.g. *β*_*Direct*_>0 in 85.0% of the posterior samples), or between antigens (e.g. *β*_*RBD*_>*β*_*N*_ in 67.2% of the posterior samples). Differences in specificity between types of assays were of epidemiologically relevant magnitude (e.g. average specificity of 99.9% (95%CrI: 99.7%-100%) RBD/Quantitative-Indirect assays, and 98.8% (95%CrI: 96.6%-99.7%) for nucleocapsid/LFA assays, see all estimates in **Table S4**). Like for sensitivity, we found considerable variability between studies reporting on the same assay (*σ*_*us*_=0.61, 95%CrI: 0.20-1.14). Specificity data and the resulting fit are shown in **Figure S7**.

### Manufacturer sensitivities and risk of bias in the literature

Although quantitatively estimating and correcting the seroreversion bias in the literature is outside the scope of the present work, we can coarsely estimate the risk of seroreversion across the literature.

First, we compared our estimates to assay sensitivities provided by manufacturers (if manufacturer values were missing, we used values reported by the FDA, or reported by authors). We found that 4 months after diagnosis, 20% of the assays have sensitivities below 75% of the originally specified value. At 6 months after diagnosis, 34% of the assays are below 75%. Thus, a few months after a COVID-19 wave, some serological assays (mostly LFA and Quantitative-Indirect assays targeting nucleocapsid antibodies) can severely underestimate previous infections.

Then, we analysed what percentage of serosurveys reported in the literature are at high risk of bias by seroreversion. As a reference, we used a comprehensive meta-analysis of the global evolution of SARS-CoV-2 seroprevalence (19), using the publicly available SeroTracker dataset (35), which notes the lack of seroreversion adjustment as a limitation. We estimated what percentage of the data points listed in SeroTracker, aligned with the WHO Unity protocol (i.e. those studies used in (19)), used assays with high rates of seroreversion (LFA assays, or nucleocapsid Quantitative-Indirect assays). Because seroreversion depends on the assay used and on the time elapsed between an epidemic wave and serosurvey, we segregated the data across semesters.

We see in **Table 2** that although the use of serological assays at high risk of seroreversion decreased throughout the pandemic, they still constituted a considerable fraction of Unity-aligned data points until mid 2021.

**Table 2.** Number of Unity-aligned seroprevalence data points of the Serotracker dataset (35) that use assays at high risk of seroreversion, defined as LFA assays or Quantitative-Indirect assays for nucleocapsid antibodies.

| Period of serological sampling | SeroTracker data points at high seroreversion risk (total data points) | Percentage of assays at high seroreversion risk (%) |
| --- | --- | --- |
| 01/01/2020 - 30/06/2020 | 135 (506) | 26.9% |
| 01/07/2020 - 31/12/2020 | 140 (596) | 23.4% |
| 01/01/2021 - 30/06/2021 | 58 (330) | 17.5% |
| 01/07/2021 - 31/12/2021 | 10 (160) | 6.3% |

## Discussion

Serology based estimates of infections are important to understand COVID-19. Although it is known that accounting for seroreversion in these estimates is important, there is a lack of appropriate data and guidelines to do so. Few studies correct for seroreversion (1,15,16,29,36,37), and the lack of robust assay-specific seroreversion estimates make it uncertain how accurate existing adjustments are. We present the first large-scale systematic analysis of seroreversion across dozens of seroassays for SARS-CoV-2, making three major contributions to help understand and correct for seroreversion.

First, we provide time-varying sensitivity estimates for 50 assays, and estimates of the average time-varying sensitivity for different assays types. These estimates can then be used to adjust for seroreversion in the literature. Knowing the assay identity (or its characteristics), and the time span between the epidemic wave and the serosurvey date at the tested location (e.g. which can be estimated from case curves), a seroreversion-adjusted sensitivity estimate can be selected from our results. Using these sensitivity estimates in the standard Gladen-Rogan formula will produce seroprevalence estimates that are corrected for seroreversion. Importantly, this procedure showed good performance at predicting assay sensitivity in a rigorous cross-validation analysis.

Our second contribution is the quantification of how seroreversion depends on assay characteristics. We show that seroreversion depends heavily on the antigen and on the analytical technology. Assays that use LFA technique (qualitative, rapid tests) show faster sensitivity decay, while quantitative assays with direct antibody binding have the slowest decay. This is in line with the high sensitivity of direct-binding assays reported for other pathogens, ascribed to factors such as less sample diluting, or the detection method not being limited to one class of antibodies (38,39). Then, assays for nucleocapsid-targeting antibodies tend to decay faster than assays for spike protein antibodies, while assays targeting S1-RBD antibodies tend to decay more slowly (although this last effect was not significant at the 95% level).

The striking differences between types of assays (e.g. average sensitivity at 6 months of 98% for S1-RBD-targeting Quantitative-Direct, against 26% for nucleocapsid LFA assays) outlines the need for assay-specific corrections. For example two seroreversion-adjusted analyses of the infection fatality-rate of SARS-CoV-2 use a single seroreversion estimate (5% monthly decrease (37), and 190 days half-life (1)) are appropriate for some assays, but can both considerably overestimate and underestimate seroreversion for other assays. These results are in line with previous reports in the literature (13–15,22), although previous studies analysed fewer characteristics in general, and did not quantify their effects. Our analysis also showed that specificity also depends on assay characteristics.

These results will allow researchers to assess the risk of seroreversion bias in serosurveys, providing a valuable tool for the design of serological studies. For example, our results suggest that the strategy of comparing S1-RBD and nucleocapsid antibody prevalences to distinguish vaccine and infection induced population immunity (10,19,40) can be affected by the different seroreversion rates of these assays.

Our third contribution is showing that a few months after diagnosis, manufacturer specifications can be highly unreliable for a considerable fraction of emergency approved assays. Relatedly, we show that a sizable fraction of Unity-aligned serosurveys used in recent WHO estimates of global seroprevalence dynamics (19) are at risk of seroreversion bias. This underscores the potential of decaying sensitivity to bias our epidemiological understanding of COVID-19, and a potential interest of public health policy makers in ensuring that assay manufacturers and regulatory organisms provide information and guidelines regarding seroreversion (41). The sensitivity estimates presented here should provide a straightforward way to correct for seroreversion in such datasets, and to quantificate literature bias.

To our knowledge, this is the most comprehensive analysis, for any pathogen, of assay-specific serological sensitivity decay and its dependence on assay characteristics. This is because some characteristics of the SARS-CoV-2 pandemic have allowed for a richer seroreversion dataset than is probably possible for any other pathogen (i.e. well approximated infection to testing times, multiple seroassays, multiple studies per seroassay, first exposures to a novel pathogen). Thus, many of the conclusions extracted from this analysis may serve as a guide for other emerging and endemic pathogens.

Our study has some limitations. First, although we included more assays than previous studies, many of the included assays counted with data for only a few time points. Second, we were unable to test the effects of important parameters such as the age or the disease severity on seroreversion (13,14,27,42). Relatedly, although an ideal dataset would use a well defined cohort, representative of the general population, with known age, sex ratio, disease severity, infecting variant, and occurrence of reinfections, the available literature falls short of this ideal. This has the potential to introduce variability and biases in our estimates. We note, however, that our modelling framework is flexible, and could be extended to account for these variables, given appropriate data. Third, as we analysed test data conditional on individuals having a previous COVID-19 diagnosis, it is likely that asymptomatic individuals are underrepresented in our sample. Fourth, because our analysis included only data points on non-vaccinated individuals, and most of the included data points were sampled in 2020 where variants of concern and reinfections were uncommon, it is unclear how our results would extrapolate to antibodies induced by vaccines, reinfections, or new variants of the virus.

## Conclusion

Accounting for seroreversion in serology-based estimates of infection numbers is important for understanding the COVID-19 pandemic, and for the usefulness of the continued use of serological testing to monitor the effects of COVID-19. Rapid LFA tests as well as Quantitative-Indirect tests for nucleocapsid targeting antibodies have a high potential for seroreversion, and Quantitative-Direct assays are likely to be preferred for long term serological surveillance. A considerable number of studies in the literature use assays with high risk of seroreversion, indicating some important potential for bias. We present a simple method for researchers to account for seroreversion when analysing serological data and when designing serological studies. This may be of interest to the management of other pathogens, and serosurveillance more in general, because of the unique opportunity to study the effects of seroreversion provided by the data generated during the COVID-19 pandemic.

## Supporting information

Supplementary methods and analysis

## Data Availability

All data is available at the associated GitHub page.

https://github.com/dherrera1911/seroreversion_metaanalysis

## Funding and competing interests

No specific funding or grant was used for this study. Authors declare no competing interests.

## Ethics statement

This study exclusively used publicly available aggregate data sets and published research, and hence no ethics approval was required.

## Notes

### Competing Interest Statement

The authors have declared no competing interest.

### Funding Statement

This study did not receive specific funding from any source.

### Author Declarations

All the data used in this manuscript were openly available in previously published studies and official reports. Links to the original data sources are found in the associated GitHub page.

### Summary of Updates

We add new analyses of the robustness of our results, we add an analysis of assay specificity, and an estimation of risk of bias in the serological literature due to seroreversion.

