## Supplementary methods and analysis for "Dynamics of SARS-CoV-2 seroassay sensitivity: a systematic review and modeling study"

Author affiliations:
1. Case Western Reserve University School of Medicine, Cleveland, USA
2. Centre for the Mathematical Modelling of Infectious Diseases, London School of Hygiene & Tropical Medicine, London, UK
3. University of Wollongong, Wollongong, Australia
4. Dartmouth College, Hanover, USA
5. National Bureau for Economic Research, Cambridge, USA
6. Centre for Economic Policy Research, London, United Kingdom
7. University of Pennsylvania, Philadelphia, PA, USA
8. Universidad de la República, Uruguay

### These authors contributed equally to the work

**Index:**

A) **Supplementary methods, literature review……………………………….. p.2**
B) **Supplementary methods, statistical modelling ………………………… p.24**
C) **Assay-specific sensitivity profiles ……………………………..…….. p.26**
D) **Assay sensitivity increase can continue across months …….………. p.37**
E) **Robustness to model architecture ………………………………………. p.38**
F) **Robustness to time estimation ..…………………………………………. p.41**
G) **Effect of assay characteristics on specificity…………………………….. p.43**

#### A) Supplementary methods, literature review

Inclusion and exclusion criteria of the systematic review

We set the following list of inclusion requirements for every study:

- Serologically tested unvaccinated individuals at least one month after a non-serological COVID-19 diagnosis
- Specified a median month of sampling, or both a sampling start-month and end-month
- Reported either confidence intervals for sensitivity, or both the number of people tested and the number of positive tests
- Used at least one commercial assay, or an assay used for seroprevalence studies (e.g. excluding studies reporting on in-house assays used only for clinical testing)

We set the following list of exclusion criteria:

- The tested population was a subset of the tested population from another included study
- Only reported the results of multiple simultaneously used assays instead of reporting assay results individually.
- Only tested sample frames likely to exclude particular age groups and other demographics (e.g. nursing home residents, transplant recipients, blood donors, children, healthcare workers, and workplace cohorts that excluded children; see (1) for further discussion)
- Only tested hospitalised patients (see (2) for further discussion)
- Used only the Wondfo or Menarini assays, which have been reported to have large between-batch variability (3,4)
- Were performed in Russia and had unknown times between diagnosis and serological sampling, since Russia's case reporting does not allow for reliable estimate of these times (see (5) for an analysis of Russia's case reporting).

The detailed list of studies and review results are available at the associated GitHub page. The review and review protocol were not registered.

Assay characteristics table

In **Table S1** we show the list of assays included in the study, indicating the main characteristics of each one, as well as the sources from which the data of each assay was obtained.

**Table S1.** List of included assays, together with data sources and assay characteristics. Note that we only indicate the “best” antigen for each assay. That is, if an assay detects antibodies targeting the nucleocapsid and the spike protein, it will appear as “Spike” in the table.

| **Assay** | **References** | **Antigen** | **Design** |
|---|---|---|---|
| Abbott Alinity | (6,7) | Nucleocapsid | Quantitative-Indirect |
| Abbott Alinity II Quant | (8) | Receptor-binding domain | Quantitative-Indirect |
| Abbott Architect | (2,6,8–26) | Nucleocapsid | Quantitative-Indirect |
| AMP rapid test | (27) | Unknown | LFA |
| Abingdon AbC-19 | (4) | Spike | LFA |
| Beckman ACCESS | (28) | Receptor-binding domain | Quantitative-Indirect |
| Biomerica rapid test | (4) | Nucleocapsid | LFA |
| BioRad Platelia | (29) | Nucleocapsid | Quantitative-Direct |
| Biozek rapid test | (4) | Receptor-binding domain | LFA |
| CCAHS | (30) | Receptor-binding domain | Quantitative-Indirect |
| COVIDAR | (31–33) | Receptor-binding domain | Quantitative-Indirect |
| Diasorin Liaison | (7,9–11,22,24,25,34) | Spike | Quantitative-Indirect |
| EDI Novel Coronavirus | (35) | Nucleocapsid | Quantitative-Indirect |
| Elecsys anti-N | (2,6,7,9–11,24,34–44) | Nucleocapsid | Quantitative-Direct |
| Elecsys anti-S | (10) | Receptor-binding domain | Quantitative-Direct |
| ERBALISA | (45,46) | Spike | Quantitative-Indirect |
| Euroimmun anti-S | (2,7,10,12,22,23,25,35,47–58) | Spike | Quantitative-Indirect |
| Euroimmun anti-N | (10,23) | Nucleocapsid | Quantitative-Indirect |
| Fortress Diagnostics | (59–61) | Spike | LFA |
| GenScript cPass | (8,10,23) | Receptor-binding domain | Quantitative-Competitive |
| Hangzhou Clongene | (62) | Unknown | LFA |
| IDK (Immundiagnostik) | (7) | Spike | Quantitative-Indirect |
| InBios | (35) | Spike | Quantitative-Indirect |
| Mehdi, in-house | (21) | Spike | Quantitative-Indirect |
| THL Finland, in-house FMIA | (63) | Spike | Quantitative-Indirect |
| THL Finland, in-house neutralisation assay | (63–65) | Receptor-binding domain | Quantitative-Competitive |

| | | | |
|---|---|---|---|
| Gauteng, in-house Luminex | (66) | Receptor-binding domain | Quantitative-Unknown |
| Tunis, in-house | (67) | Receptor-binding domain | Quantitative-Unknown |
| Kavach ELISA IgG (Zydus) | (45) | Receptor-binding domain | Quantitative-Indirect |
| Livzon lateral flow | (68,69) | Unknown | LFA |
| LumiraDx | (4) | Receptor-binding domain | Quantitative-Unknown |
| Luminex xMAP (RBD portion) | (70) | Receptor-binding domain | Quantitative-Indirect |
| Massachusetts General Hospital | (71) | Receptor-binding domain | Quantitative-Indirect |
| Mount Sinai (New York) | (72) | Receptor-binding domain | Quantitative-Indirect |
| Roche rapid test | (4) | Spike | LFA |
| Orient Gene Biotech | (13,73,74) | Spike | LFA |
| SenASTrIS test | (75,76) | Spike | Quantitative-Indirect |
| Siemens Advia Centaur | (24,77) | Receptor-binding domain | Quantitative-Direct |
| Siemens Advia anti-RBD | (20) | Receptor-binding domain | Quantitative-Indirect |
| Standard Q | (78) | Receptor-binding domain | LFA |
| Sinai Health (Toronto) | (79) | Spike | Quantitative-Indirect |
| Techno Genetics | (80) | Receptor-binding domain | LFA |
| Tosoh N sandwich | (43) | Nucleocapsid | Quantitative-Direct |
| Tosoh N indirect | (43) | Nucleocapsid | Quantitative-Indirect |
| Tosoh S sandwich | (43) | Receptor-binding domain | Quantitative-Direct |
| Tosoh S indirect | (43) | Receptor-binding domain | Quantitative-Indirect |
| University of Rio de Janeiro | (3,53) | Spike | Quantitative-Indirect |
| Vitros Ortho (not total antibodies) | (9,81–84) | Spike | Quantitative-Indirect |
| Vitros Ortho (total antibodies) | (9,10) | Receptor-binding domain | Quantitative-Direct |
| Wantai | (10,42,85–88) | Receptor-binding domain | Quantitative-Direct |

Data extraction and risk of bias assessment

Two authors reviewed the studies, screened for inclusion, extracted data, and assessed studies for risk of bias. All disagreements were solved by consensus via email and bi-monthly meetings. Where essential data were missing, we requested the data from study authors by email.

We extracted the following data for each included study:
● Nation or subnational location from which the sample was drawn
● Sample frame
● Median month of sampling, and/or both a sampling start-month and end-month
● Either confidence intervals for sensitivity, and/or both the number of people tested and the number of positive tests
● Name of the serological assay
● Reported median (or mean) age and sex balance of the cohort
● Reported severity of infection in the cohort

For each included assay we sought reported sensitivity from the manufacturer or other published source.

We assessed studies as low risk of bias if participants were drawn from a random sample taken in a sample frame representative of the general population, or from sampling the majority of the population by census. Detailed assessment comments are available in the associated GitHub page (Methods section, main text). Studies were assessed as high risk when participants were drawn from an alternative sample, such as a convenience sample of non-hospitalized individuals. Publication bias is unlikely to strongly skew our results because studies with high or low seropositivity rates are of scientific interest; prior work found no evidence of publication bias in seroprevalence studies within wealthier nations (85). We sought to further minimise risk of publication bias by extensively searching the grey literature and following up on media reports (86).

In **Table S2**, we show a list of the included studies and each data point included in our analysis, indicating different relevant cohort characteristics. We observe that the median age of the studies falls mostly in the range around 40 years, as one would expect from representative samples of the adult population, with some somewhat older and some younger cohorts. The sex ratio in the samples is also mostly centred around 50% female participants, with some studies reporting somewhat unbalanced samples (e.g. 70% female, 30% male), suggesting that some studies depart from the expected representative ratios (although some cohorts are small, and random variability in sex ratio can be expected to be large). We also show the infection severity of the cohorts when these are reported. The majority of studies reporting the severity of the cohort have severity ranges under 10%, which is in the range expected from the rate of severe disease among diagnosed individuals in 2020 (89), when most studies were performed. Some studies report higher hospitalisation rates, indicating that these samples are not representative of the general population. It is likely that most of the studies not reporting on the rate of severe disease include only small numbers of patients with severe disease (i.e. they are not sampling a cohort where hospitalised patients are overrepresented and where reporting on severity would be crucial). Under this assumption, most of included studies have rates of severe disease within the expected range. In sum, the reported ages, sex ratios and severity rates of analyzed cohorts indicate that most data points are broadly in line with the expectations of a representative population sampling. Some cohorts depart from the expected representative numbers, indicating a potential to bias the obtained results. However, because most data points have apparently representative cohorts, and non-representative cohorts are distributed among the different types of tests, we expect the potential for bias from cohort characteristics to be small.

Two further concerns regarding potential bias in the results are the effects of infection by different variants, as well as the potential for reinfections. No studies report on the infecting variant of the tested individuals, and only one study reports on the absence of reinfections (22). However, both the emergence of new SARS-CoV-2 variants and reinfections were not common during most of 2020, and so we assess that the serology samples collected in 2020 are likely due to infections from the original variant, and at low risk of reinfections. We see in **Table S2** that most analysed samples (90%, 217/242 data points with reported sampling date) were collected in 2020, suggesting that our results are mostly representative of the original variant, and at low risk of bias due to reinfections.

**Table S2.** Characteristics of serological studies and analysed cohorts for each data point included in the study. Empty cells indicate that the value is not reported in the study. For rows with empty time between diagnosis and serology, we estimated the times as reported in the Methods section in the main text. Each row corresponds to one data point in **Figure S1** below.

| Reference | Serology sampling start date | Serology sampling end date | Serology sampling midpoint | Time between prior diagnosis (or symptoms) and serology (Months) | Assay | Severity of infection | Median age | Sex ratio | Notes |
|---|---|---|---|---|---|---|---|---|---|
| 2 | NA | October 2020 | October 2020 | 6 | Abbott Architect | 0% hospitalized | NA | NA | |
| 2 | NA | October 2020 | October 2020 | 6 | Elecsys anti-N | 0% hospitalized | NA | NA | |
| 2 | NA | October 2020 | October 2020 | 6 | Euroimmun anti-S | 0% hospitalized | NA | NA | |
| 3 | NA | NA | November 2020 | 1 | University of Rio de Janeiro | NA | 30 - 50 | 58% female , 42% male | table 2 |
| 3 | NA | NA | November 2020 | 2 | University of Rio de Janeiro | NA | 30 - 50 | 58% female , 42% male | table 2 |
| 3 | NA | NA | November 2020 | 3 | University of Rio de Janeiro | NA | 30 - 50 | 58% female , 42% male | table 2 |
| 3 | NA | NA | November 2020 | 4 | University of Rio de Janeiro | NA | 30 - 50 | 58% female , 42% male | table 2 |
| 3 | NA | NA | November 2020 | 5 | University of Rio de Janeiro | NA | 30 - 50 | 58% female , 42% male | table 2 |
| 4 | September 2020 | November 2020 | NA | 2 | Abingdon AbC-19 | 2.6% hospitalized | NA | NA | |
| 4 | September 2020 | November 2020 | NA | 4 | Abingdon AbC-19 | 2.6% hospitalized | NA | NA | |
| 4 | September 2020 | November 2020 | NA | 6 | Abingdon AbC-19 | 2.6% hospitalized | NA | NA | |
| 4 | September 2020 | November 2020 | NA | 1 | Biomerica rapid test | 2.6% hospitalized | NA | NA | |
| 4 | September 2020 | November 2020 | NA | 2 | Biomerica rapid test | 2.6% hospitalized | NA | NA | |

| | | | | | | | | | |
|---|---|---|---|---|---|---|---|---|---|
| 4 | September 2020 | November 2020 | NA | 4 | Biomerica rapid test | 2.6% hospitalized | NA | NA | |
| 4 | September 2020 | November 2020 | NA | 6 | Biomerica rapid test | 2.6% hospitalized | NA | NA | |
| 4 | September 2020 | November 2020 | NA | 1 | Biozek rapid test | 2.6% hospitalized | NA | NA | |
| 4 | September 2020 | November 2020 | NA | 2 | Biozek rapid test | 2.6% hospitalized | NA | NA | |
| 4 | September 2020 | November 2020 | NA | 1 | LumiraDx | 2.6% hospitalized | NA | NA | |
| 4 | September 2020 | November 2020 | NA | 2 | LumiraDx | 2.6% hospitalized | NA | NA | |
| 4 | September 2020 | November 2020 | NA | 4 | LumiraDx | 2.6% hospitalized | NA | NA | |
| 4 | September 2020 | November 2020 | NA | 6 | LumiraDx | 2.6% hospitalized | NA | NA | |
| 4 | September 2020 | November 2020 | NA | 1 | Roche rapid test | 2.6% hospitalized | NA | NA | |
| 4 | September 2020 | November 2020 | NA | 2 | Roche rapid test | 2.6% hospitalized | NA | NA | |
| 4 | September 2020 | November 2020 | NA | 4 | Roche rapid test | 2.6% hospitalized | NA | NA | |
| 4 | September 2020 | November 2020 | NA | 6 | Roche rapid test | 2.6% hospitalized | NA | NA | |
| 6 | 3/26/2020 | 7/10/2020 | 5/18/2020 | 1 | Abbott Alinity | 72% hospitalized | 57 | 46% female , 54% male | table 1 |
| 6 | 3/26/2020 | 7/10/2020 | 5/18/2020 | 1 | Abbott Architect | 72% hospitalized | 57 | 46% female , 54% male | table 1 |
| 6 | 3/26/2020 | 7/10/2020 | 5/18/2020 | 1 | Elecsys anti-N | 72% hospitalized | 57 | 46% female , 54% male | table 1 |
| 7 | August 2020 | September 2020 | NA | 5 | Abbott Alinity | 4% severe | 44 | 55% female , 45% male | table 2 |

| | | | | | | | | | |
|---|---|---|---|---|---|---|---|---|---|
| 7 | August 2020 | September 2020 | NA | 5 | Diasorin Liaison | 4% severe | 44 | 55% female , 45% male | table 2 |
| 7 | August 2020 | September 2020 | NA | 5 | Elecsys anti-N | 4% severe | 44 | 55% female , 45% male | table 2 |
| 7 | August 2020 | September 2020 | NA | 5 | Euroimmun anti-S | 4% severe | 44 | 55% female , 45% male | table 2 |
| 7 | August 2020 | September 2020 | NA | 5 | IDK (Immundiagnostik) | 4% severe | 44 | 55% female , 45% male | table 2 |
| 8 | 3/11/2020 | 4/5/2020 | NA | 10 | Abbott Alinity II Quant | NA | 52 | 70% female , 30% male | table 1 |
| 8 | 3/11/2020 | 4/5/2020 | NA | 10 | Abbott Architect | NA | 52 | 70% female , 30% male | table 1 |
| 8 | 3/11/2020 | 4/5/2020 | NA | 10 | GenScript cPass | NA | 52 | 70% female , 30% male | table 1 |
| 9 | April 2020 | July 2020 | NA | 1 | Abbott Architect | 0% hospitalized | 30 - 45 | 48% female , 52% male | supplementary table 1 |
| 9 | NA | NA | NA | 3 | Abbott Architect | 0% hospitalized | 30 - 45 | 48% female , 52% male | supplementary table 1 |
| 9 | NA | NA | NA | 5 | Abbott Architect | 0% hospitalized | 30 - 45 | 48% female , 52% male | supplementary table 1 |
| 9 | NA | NA | NA | 7 | Abbott Architect | 0% hospitalized | 30 - 45 | 48% female , 52% male | supplementary table 1 |
| 9 | April 2020 | July 2020 | NA | 1 | Diasorin Liaison | 0% hospitalized | 30 - 45 | 48% female , 52% male | supplementary table 1 |
| 9 | NA | NA | NA | 3 | Diasorin Liaison | 0% hospitalized | 30 - 45 | 48% female , 52% male | supplementary table 1 |
| 9 | NA | NA | NA | 5 | Diasorin Liaison | 0% hospitalized | 30 - 45 | 48% female , 52% male | supplementary table 1 |

| | | | | | | | | | |
|---|---|---|---|---|---|---|---|---|---|
| 9 | NA | NA | NA | 7 | Diasorin Liaison | 0% hospitalized | 30 - 45 | 48% female , 52% male | supplementary table 1 |
| 9 | April 2020 | July 2020 | NA | 1 | Elecsys anti-N | 0% hospitalized | 30 - 45 | 48% female , 52% male | supplementary table 1 |
| 9 | NA | NA | NA | 3 | Elecsys anti-N | 0% hospitalized | 30 - 45 | 48% female , 52% male | supplementary table 1 |
| 9 | NA | NA | NA | 5 | Elecsys anti-N | 0% hospitalized | 30 - 45 | 48% female , 52% male | supplementary table 1 |
| 9 | NA | NA | NA | 7 | Elecsys anti-N | 0% hospitalized | 30 - 45 | 48% female , 52% male | supplementary table 1 |
| 9 | April 2020 | July 2020 | NA | 1 | Vitros Ortho (not total antibodies) | 0% hospitalized | 30 - 45 | 48% female , 52% male | supplementary table 1 |
| 9 | NA | NA | NA | 3 | Vitros Ortho (not total antibodies) | 0% hospitalized | 30 - 45 | 48% female , 52% male | supplementary table 1 |
| 9 | NA | NA | NA | 5 | Vitros Ortho (not total antibodies) | 0% hospitalized | 30 - 45 | 48% female , 52% male | supplementary table 1 |
| 9 | NA | NA | NA | 7 | Vitros Ortho (not total antibodies) | 0% hospitalized | 30 - 45 | 48% female , 52% male | supplementary table 1 |
| 9 | April 2020 | July 2020 | NA | 1 | Vitros Ortho (total antibodies) | 0% hospitalized | 30 - 45 | 48% female , 52% male | supplementary table 1 |
| 9 | NA | NA | NA | 3 | Vitros Ortho (total antibodies) | 0% hospitalized | 30 - 45 | 48% female , 52% male | supplementary table 1 |
| 9 | NA | NA | NA | 5 | Vitros Ortho (total antibodies) | 0% hospitalized | 30 - 45 | 48% female , 52% male | supplementary table 1 |
| 9 | NA | NA | NA | 7 | Vitros Ortho (total antibodies) | 0% hospitalized | 30 - 45 | 48% female , 52% male | supplementary table 1 |
| 10 | NA | NA | 2020 | 1 | Abbott Architect | 15.8% hospitalized | NA | NA | |
| 10 | NA | NA | 2020 | 2 | Abbott Architect | 15.8% hospitalized | NA | NA | |
| 10 | NA | NA | 2020 | 4 | Abbott Architect | 15.8% hospitalized | NA | NA | |

| | | | | | | | | |
|---|---|---|---|---|---|---|---|---|
| 10 | NA | NA | 2020 | 6 | Abbott Architect | 15.8% hospitalized | NA | NA |
| 10 | NA | NA | 2020 | 8 | Abbott Architect | 15.8% hospitalized | NA | NA |
| 10 | NA | NA | 2020 | 10 | Abbott Architect | 15.8% hospitalized | NA | NA |
| 10 | NA | NA | 2020 | 12 | Abbott Architect | 15.8% hospitalized | NA | NA |
| 10 | NA | NA | 2020 | 14 | Abbott Architect | 15.8% hospitalized | NA | NA |
| 10 | NA | NA | 2020 | 1 | Diasorin Liaison | 15.8% hospitalized | NA | NA |
| 10 | NA | NA | 2020 | 2 | Diasorin Liaison | 15.8% hospitalized | NA | NA |
| 10 | NA | NA | 2020 | 4 | Diasorin Liaison | 15.8% hospitalized | NA | NA |
| 10 | NA | NA | 2020 | 6 | Diasorin Liaison | 15.8% hospitalized | NA | NA |
| 10 | NA | NA | 2020 | 8 | Diasorin Liaison | 15.8% hospitalized | NA | NA |
| 10 | NA | NA | 2020 | 10 | Diasorin Liaison | 15.8% hospitalized | NA | NA |
| 10 | NA | NA | 2020 | 12 | Diasorin Liaison | 15.8% hospitalized | NA | NA |
| 10 | NA | NA | 2020 | 14 | Diasorin Liaison | 15.8% hospitalized | NA | NA |
| 10 | NA | NA | 2020 | 1 | Elecsys anti-N | 15.8% hospitalized | NA | NA |
| 10 | NA | NA | 2020 | 2 | Elecsys anti-N | 15.8% hospitalized | NA | NA |
| 10 | NA | NA | 2020 | 4 | Elecsys anti-N | 15.8% hospitalized | NA | NA |

| | | | | | | | | |
|---|---|---|---|---|---|---|---|---|
| 10 | NA | NA | 2020 | 6 | Elecsys anti-N | 15.8% hospitalized | NA | NA |
| 10 | NA | NA | 2020 | 8 | Elecsys anti-N | 15.8% hospitalized | NA | NA |
| 10 | NA | NA | 2020 | 10 | Elecsys anti-N | 15.8% hospitalized | NA | NA |
| 10 | NA | NA | 2020 | 12 | Elecsys anti-N | 15.8% hospitalized | NA | NA |
| 10 | NA | NA | 2020 | 14 | Elecsys anti-N | 15.8% hospitalized | NA | NA |
| 10 | NA | NA | 2020 | 1 | Elecsys anti-S | 15.8% hospitalized | NA | NA |
| 10 | NA | NA | 2020 | 2 | Elecsys anti-S | 15.8% hospitalized | NA | NA |
| 10 | NA | NA | 2020 | 4 | Elecsys anti-S | 15.8% hospitalized | NA | NA |
| 10 | NA | NA | 2020 | 6 | Elecsys anti-S | 15.8% hospitalized | NA | NA |
| 10 | NA | NA | 2020 | 8 | Elecsys anti-S | 15.8% hospitalized | NA | NA |
| 10 | NA | NA | 2020 | 10 | Elecsys anti-S | 15.8% hospitalized | NA | NA |
| 10 | NA | NA | 2020 | 12 | Elecsys anti-S | 15.8% hospitalized | NA | NA |
| 10 | NA | NA | 2020 | 14 | Elecsys anti-S | 15.8% hospitalized | NA | NA |
| 10 | NA | NA | 2020 | 1 | Euroimmun anti-N | 15.8% hospitalized | NA | NA |
| 10 | NA | NA | 2020 | 2 | Euroimmun anti-N | 15.8% hospitalized | NA | NA |
| 10 | NA | NA | 2020 | 4 | Euroimmun anti-N | 15.8% hospitalized | NA | NA |

| | | | | | | | | |
|---|---|---|---|---|---|---|---|---|
| 10 | NA | NA | 2020 | 6 | Euroimmun anti-N | 15.8% hospitalized | NA | NA |
| 10 | NA | NA | 2020 | 8 | Euroimmun anti-N | 15.8% hospitalized | NA | NA |
| 10 | NA | NA | 2020 | 10 | Euroimmun anti-N | 15.8% hospitalized | NA | NA |
| 10 | NA | NA | 2020 | 12 | Euroimmun anti-N | 15.8% hospitalized | NA | NA |
| 10 | NA | NA | 2020 | 14 | Euroimmun anti-N | 15.8% hospitalized | NA | NA |
| 10 | NA | NA | 2020 | 1 | Euroimmun anti-S | 15.8% hospitalized | NA | NA |
| 10 | NA | NA | 2020 | 2 | Euroimmun anti-S | 15.8% hospitalized | NA | NA |
| 10 | NA | NA | 2020 | 4 | Euroimmun anti-S | 15.8% hospitalized | NA | NA |
| 10 | NA | NA | 2020 | 6 | Euroimmun anti-S | 15.8% hospitalized | NA | NA |
| 10 | NA | NA | 2020 | 8 | Euroimmun anti-S | 15.8% hospitalized | NA | NA |
| 10 | NA | NA | 2020 | 10 | Euroimmun anti-S | 15.8% hospitalized | NA | NA |
| 10 | NA | NA | 2020 | 12 | Euroimmun anti-S | 15.8% hospitalized | NA | NA |
| 10 | NA | NA | 2020 | 14 | Euroimmun anti-S | 15.8% hospitalized | NA | NA |
| 10 | NA | NA | 2020 | 1 | GenScript cPass | 15.8% hospitalized | NA | NA |
| 10 | NA | NA | 2020 | 2 | GenScript cPass | 15.8% hospitalized | NA | NA |
| 10 | NA | NA | 2020 | 4 | GenScript cPass | 15.8% hospitalized | NA | NA |

| | | | | | | | | |
|---|---|---|---|---|---|---|---|---|
| 10 | NA | NA | 2020 | 6 | GenScript cPass | 15.8% hospitalized | NA | NA |
| 10 | NA | NA | 2020 | 8 | GenScript cPass | 15.8% hospitalized | NA | NA |
| 10 | NA | NA | 2020 | 10 | GenScript cPass | 15.8% hospitalized | NA | NA |
| 10 | NA | NA | 2020 | 12 | GenScript cPass | 15.8% hospitalized | NA | NA |
| 10 | NA | NA | 2020 | 14 | GenScript cPass | 15.8% hospitalized | NA | NA |
| 10 | NA | NA | 2020 | 1 | Vitros Ortho (total antibodies) | 15.8% hospitalized | NA | NA |
| 10 | NA | NA | 2020 | 2 | Vitros Ortho (total antibodies) | 15.8% hospitalized | NA | NA |
| 10 | NA | NA | 2020 | 4 | Vitros Ortho (total antibodies) | 15.8% hospitalized | NA | NA |
| 10 | NA | NA | 2020 | 6 | Vitros Ortho (total antibodies) | 15.8% hospitalized | NA | NA |
| 10 | NA | NA | 2020 | 8 | Vitros Ortho (total antibodies) | 15.8% hospitalized | NA | NA |
| 10 | NA | NA | 2020 | 10 | Vitros Ortho (total antibodies) | 15.8% hospitalized | NA | NA |
| 10 | NA | NA | 2020 | 12 | Vitros Ortho (total antibodies) | 15.8% hospitalized | NA | NA |
| 10 | NA | NA | 2020 | 14 | Vitros Ortho (total antibodies) | 15.8% hospitalized | NA | NA |
| 10 | NA | NA | 2020 | 1 | Wantai | 15.8% hospitalized | NA | NA |
| 10 | NA | NA | 2020 | 2 | Wantai | 15.8% hospitalized | NA | NA |
| 10 | NA | NA | 2020 | 4 | Wantai | 15.8% hospitalized | NA | NA |

| | | | | | | | | | |
|---|---|---|---|---|---|---|---|---|---|
| 10 | NA | NA | 2020 | 6 | Wantai | 15.8% hospitalized | NA | NA | |
| 10 | NA | NA | 2020 | 8 | Wantai | 15.8% hospitalized | NA | NA | |
| 10 | NA | NA | 2020 | 10 | Wantai | 15.8% hospitalized | NA | NA | |
| 10 | NA | NA | 2020 | 12 | Wantai | 15.8% hospitalized | NA | NA | |
| 10 | NA | NA | 2020 | 14 | Wantai | 15.8% hospitalized | NA | NA | |
| 11 | NA | NA | NA | 1 | Abbott Architect | ~26% hospitalized | 40 - 45 | ~42% female , ~58% male | supplementary data #2 |
| 11 | NA | NA | NA | 1 - 3 | Abbott Architect | ~26% hospitalized | 40 - 45 | ~42% female , ~58% male | supplementary data #2 |
| 11 | NA | NA | NA | 4 - 5 | Abbott Architect | ~26% hospitalized | 40 - 45 | ~42% female , ~58% male | supplementary data #2 |
| 11 | NA | NA | NA | 5 - 7 | Abbott Architect | ~26% hospitalized | 40 - 45 | ~42% female , ~58% male | supplementary data #2 |
| 11 | NA | NA | NA | 1 | Diasorin Liaison | ~26% hospitalized | 40 - 45 | ~42% female , ~58% male | supplementary data #2 |
| 11 | NA | NA | NA | 1 - 3 | Diasorin Liaison | ~26% hospitalized | 40 - 45 | ~42% female , ~58% male | supplementary data #2 |
| 11 | NA | NA | NA | 4 - 5 | Diasorin Liaison | ~26% hospitalized | 40 - 45 | ~42% female , ~58% male | supplementary data #2 |
| 11 | NA | NA | NA | 5 - 7 | Diasorin Liaison | ~26% hospitalized | 40 - 45 | ~42% female , ~58% male | supplementary data #2 |
| 11 | NA | NA | NA | 1 | Elecsys anti-N | ~26% hospitalized | 40 - 45 | ~42% female , ~58% male | supplementary data #2 |
| 11 | NA | NA | NA | 1 - 3 | Elecsys anti-N | ~26% hospitalized | 40 - 45 | ~42% female , ~58% male | supplementary data #2 |
| 11 | NA | NA | NA | 4 - 5 | Elecsys anti-N | ~26% hospitalized | 40 - 45 | ~42% female , ~58% male | supplementary data #2 |

| | | | | | | | | | |
|---|---|---|---|---|---|---|---|---|---|
| 11 | NA | NA | NA | 5 - 7 | Elecsys anti-N | ~26% hospitalized | 40 - 45 | ~42% female , ~58% male | supplementary data #2 |
| 12 | 4/21/2020 | 4/27/2020 | 4/24/2020 | NA | Abbott Architect | < 9% hospitalized | 30 - 50 | ~50% | supplementary figure 5B |
| 12 | 4/21/2020 | 4/27/2020 | 4/24/2020 | NA | Euroimmun anti-S | < 9% hospitalized | 30 - 50 | ~50% | supplementary figure 5B |
| 13 | 4/27/2020 | 5/11/2020 | 5/4/2020 | NA | Abbott Architect | NA | NA | NA | |
| 13 | 4/27/2020 | 5/11/2020 | 5/4/2020 | NA | Orient Gene Biotech | NA | NA | NA | |
| 14 | 5/1/2020 | 6/30/2020 | 5/31/2020 | NA | Abbott Architect | NA | NA | NA | |
| 15 | 6/24/2020 | 7/8/2020 | 7/1/2020 | NA | Abbott Architect | NA | NA | NA | |
| 16 | 8/17/2020 | 9/22/2020 | 9/4/2020 | NA | Abbott Architect | NA | NA | NA | |
| 17 | 10/17/2020 | 10/20/2020 | 10/18/2020 | NA | Abbott Architect | NA | NA | NA | |
| 18 | 10/17/2020 | 11/4/2020 | 10/26/2020 | NA | Abbott Architect | ~49% symptomatic | NA | NA | figure 3 |
| 19 | 6/14/2021 | 7/6/2021 | 6/25/2021 | NA | Abbott Architect | NA | NA | NA | |
| 20 | NA | NA | 12/27/2020 | 0.5 - 2 | Abbott Architect | NA | NA | NA | |
| 20 | NA | NA | 12/27/2020 | 2 - 4 | Abbott Architect | NA | NA | NA | |
| 20 | NA | NA | 12/27/2020 | 4 - 6 | Abbott Architect | NA | NA | NA | |
| 20 | NA | NA | 12/27/2020 | >6 | Abbott Architect | NA | NA | NA | |
| 20 | NA | NA | 12/27/2020 | 0.5 - 2 | Siemens Advia anti-RBD | NA | NA | NA | |
| 20 | NA | NA | 12/27/2020 | 2 - 4 | Siemens Advia anti-RBD | NA | NA | NA | |
| 20 | NA | NA | 12/27/2020 | 4 - 6 | Siemens Advia anti-RBD | NA | NA | NA | |
| 20 | NA | NA | 12/27/2020 | >6 | Siemens Advia anti-RBD | NA | NA | NA | |
| 21 | NA | NA | 12/18/2020 | 2 | Abbott Architect | 18% require oxygen | 40 | 30% female , 70% male | table 1 |
| 21 | NA | NA | 12/18/2020 | 2 | Mehdi, in-house | 18% require oxygen | 40 | 30% female , 70% male | table 1 |

| | | | | | | | | | |
|---|---|---|---|---|---|---|---|---|---|
| 22 | March 2020 | August 2020 | NA | 2 | Abbott Architect | 12% hospitalized | 48 | 58% female , 42% male | table 1 |
| 22 | March 2020 | August 2020 | NA | 2 | Diasorin Liaison | 12% hospitalized | 48 | 58% female , 42% male | table 1 |
| 22 | March 2020 | August 2020 | NA | 2 | Euroimmun anti-S | 12% hospitalized | 48 | 58% female , 42% male | table 1 |
| 23 | 4/14/2020 | NA | NA | 2 | Euroimmun anti-N | NA | NA | NA | |
| 23 | NA | 8/15/2020 | NA | 4 | Euroimmun anti-N | NA | NA | NA | |
| 23 | 4/14/2020 | NA | NA | 2 | Euroimmun anti-S | NA | NA | NA | |
| 23 | NA | 8/15/2020 | NA | 4 | Euroimmun anti-S | NA | NA | NA | |
| 23 | 4/14/2020 | NA | NA | 2 | GenScript cPass | NA | NA | NA | |
| 23 | NA | 8/15/2020 | NA | 4 | GenScript cPass | NA | NA | NA | |
| 24 | NA | NA | NA | 3 | Abbott Architect | 0% hospitalized | 44 | 72% female , 28% male | |
| 24 | NA | NA | NA | 3 | Diasorin Liaison | 0% hospitalized | 44 | 72% female , 28% male | |
| 24 | NA | NA | NA | 3 | Elecsys anti-N | 0% hospitalized | 44 | 72% female , 28% male | |
| 24 | NA | NA | NA | 3 | Siemens Advia Centaur | 0% hospitalized | 44 | 72% female , 28% male | |
| 25 | August 2020 | October 2020 | September 2020 | 5 | Abbott Architect | 7% hospitalized | 46 | 52% female , 48% male | table 1 |
| 25 | January 2021 | March 2021 | February 2021 | 10 | Abbott Architect | 7% hospitalized | 46 | 52% female , 48% male | table 1 |
| 25 | August 2020 | October 2020 | September 2020 | 5 | Diasorin Liaison | 7% hospitalized | 46 | 52% female , 48% male | table 1 |
| 25 | January 2021 | March 2021 | February 2021 | 10 | Diasorin Liaison | 7% hospitalized | 46 | 52% female , 48% male | table 1 |
| 25 | August 2020 | October 2020 | September 2020 | 5 | Euroimmun anti-S | 7% hospitalized | 46 | 52% female , 48% male | table 1 |
| 25 | January 2021 | March 2021 | February 2021 | 10 | Euroimmun anti-S | 7% hospitalized | 46 | 52% female , 48% male | table 1 |

| | | | | | | | | | |
|---|---|---|---|---|---|---|---|---|---|
| 26 | 7/17/2020 | 10/15/2020 | 8/31/2020 | 1 - 2 | Abbott Architect | NA | NA | NA | |
| 26 | 7/17/2020 | 10/15/2020 | 8/31/2020 | 2 - 3 | Abbott Architect | NA | NA | NA | |
| 26 | 7/17/2020 | 10/15/2020 | 8/31/2020 | 3 - 4 | Abbott Architect | NA | NA | NA | |
| 26 | 7/17/2020 | 10/15/2020 | 8/31/2020 | 4 - 5 | Abbott Architect | NA | NA | NA | |
| 27 | 8/10/2020 | 9/10/2020 | 8/25/2020 | NA | AMP rapid test | NA | NA | NA | email from study authors |
| 28 | 8/15/2020 | 9/5/2020 | 8/25/2020 | NA | Beckman ACCESS | 0% hospitalized | NA | NA | |
| 28 | 9/12/2020 | 10/24/2020 | 10/3/2020 | NA | Beckman ACCESS | 0% hospitalized | NA | NA | |
| 28 | 11/7/2020 | 12/19/2020 | 11/28/2020 | NA | Beckman ACCESS | 0% hospitalized | NA | NA | |
| 29 | NA | NA | 10/30/2020 | NA | BioRad Platelia | NA | NA | NA | |
| 30 | 11/2/2020 | 4/30/2021 | 1/30/2021 | NA | CCAHS | NA | NA | NA | |
| 31 | 03/03/2021 | 4/17/2021 | 03/25/2021 | NA | COVIDAR | ~4% hospitalized | 39 | 61% female , 39% male | positive on nasal swab (supplemental S2 data) |
| 32 | NA | NA | NA | 1 | COVIDAR | NA | NA | NA | |
| 33 | NA | September 2020 | NA | 1 | COVIDAR | 10% severe | 37 - 45 | 60% female , 40% male | table 1 |
| 33 | NA | September 2020 | NA | 3 | COVIDAR | 12% severe | 36 - 50 | 65% female , 35% male | table 1 |
| 34 | 5/1/2020 | 5/3/2020 | 5/2/2020 | 2 | Abbott Architect | NA | NA | NA | |
| 34 | 5/1/2020 | 5/3/2020 | 5/2/2020 | 2 | Diasorin Liaison | NA | NA | NA | |
| 34 | 5/1/2020 | 5/3/2020 | 5/2/2020 | 2 | Elecsys anti-N | NA | NA | NA | |
| 35 | NA | NA | NA | 8 | EDI Novel Coronavirus | 0% hospitalized | 26 | 60% female , 40% male | tables 1 and 2 |
| 35 | NA | NA | NA | 8 | Elecsys anti-N | 0% hospitalized | 26 | 60% female , 40% male | tables 1 and 2 |

| | | | | | | | | | |
|---|---|---|---|---|---|---|---|---|---|
| 35 | NA | NA | NA | 8 | Euroimmun anti-S | 0% hospitalized | 26 | 60% female , 40% male | tables 1 and 2 |
| 35 | NA | NA | NA | 8 | InBios | 0% hospitalized | 26 | 60% female , 40% male | tables 1 and 2 |
| 36 | 8/11/2020 | 8/16/2020 | 8/13/2020 | NA | Elecsys anti-N | NA | NA | NA | |
| 36 | 9/10/2020 | 9/16/2020 | 9/13/2020 | NA | Elecsys anti-N | NA | NA | NA | |
| 36 | 10/12/2020 | 10/16/2020 | 10/14/2020 | NA | Elecsys anti-N | NA | NA | NA | |
| 37 | 11/23/2020 | 2/19/2021 | 1/6/2021 | NA | Elecsys anti-N | NA | NA | NA | |
| 38 | 1/8/2021 | 1/24/2021 | 1/16/2021 | NA | Elecsys anti-N | NA | NA | NA | |
| 39 | 12/3/2021 | 12/27/2021 | 12/15/2021 | NA | Elecsys anti-N | NA | NA | NA | |
| 39 | 2/2/2022 | 3/6/2022 | 2/18/2022 | NA | Elecsys anti-N | NA | NA | NA | |
| 40 | 8/6/2020 | 8/31/2020 | 8/18/2020 | 1 | Elecsys anti-N | NA | NA | NA | |
| 41 | NA | NA | 8/26/2020 | 1 | Elecsys anti-N | NA | NA | NA | |
| 41 | NA | NA | 9/18/2020 | 1 | Elecsys anti-N | NA | NA | NA | |
| 42 | 4/3/2020 | 7/8/2020 | 5/21/2020 | 3 | Elecsys anti-N | 3% hospitalized | 43 | 52% female , 48% male | table S1 in appendix 1, appendix 2 |
| 42 | 4/3/2020 | 7/8/2020 | 5/21/2020 | 3 | Wantai | 3% hospitalized | 43 | 52% female , 48% male | table S1 in appendix 1, appendix 2 |
| 43 | 9/2/2020 | 10/26/2020 | 9/29/2020 | 6 | Elecsys anti-N | 73% hospitalized | 50 | 50% female , 50% male | table 1 |
| 43 | 9/2/2020 | 10/26/2020 | 9/29/2020 | 6 | Tosoh N indirect | 73% hospitalized | 50 | 50% female , 50% male | table 1 |
| 43 | 9/2/2020 | 10/26/2020 | 9/29/2020 | 6 | Tosoh N sandwich | 73% hospitalized | 50 | 50% female , 50% male | table 1 |
| 43 | 9/2/2020 | 10/26/2020 | 9/29/2020 | 6 | Tosoh S indirect | 73% hospitalized | 50 | 50% female , 50% male | table 1 |
| 43 | 9/2/2020 | 10/26/2020 | 9/29/2020 | 6 | Tosoh S sandwich | 73% hospitalized | 50 | 50% female , 50% male | table 1 |

| | | | | | | | | | |
|---|---|---|---|---|---|---|---|---|---|
| 44 | NA | late 2020 | NA | 9 | Elecsys anti-N | 3% hospitalized | 57 | 19% female , 81% male | table 2 |
| 45 | 9/1/2020 | 9/7/2020 | 9/4/2020 | NA | ERBALISA | NA | NA | NA | |
| 45 | 10/15/2020 | 10/21/2020 | 10/18/2020 | NA | ERBALISA | NA | NA | NA | |
| 45 | 8/1/2020 | 8/7/2020 | 8/4/2020 | NA | Kavach ELISA IgG (Zydus) | NA | NA | NA | |
| 46 | 7/27/2020 | 8/7/2020 | 8/1/2020 | 2 | ERBALISA | NA | NA | NA | |
| 47 | NA | NA | 5/21/2020 | NA | Euroimmun anti-S | NA | NA | NA | |
| 48 | 5/4/2020 | 6/23/2020 | 5/29/2020 | NA | Euroimmun anti-S | NA | NA | NA | |
| 49 | 5/20/2020 | 6/9/2020 | 5/30/2020 | NA | Euroimmun anti-S | 34% moderate or severe | 52 | NA | table 3 |
| 50 | 7/20/2020 | 8/5/2020 | 7/28/2020 | NA | Euroimmun anti-S | NA | NA | NA | |
| 51 | 7/1/2020 | 7/28/2020 | 7/14/2020 | NA | Euroimmun anti-S | NA | 46 - 65 | 57% female, 43% male | supplement tables 1 + 2 |
| 51 | 8/5/2020 | 9/1/2020 | 8/18/2020 | NA | Euroimmun anti-S | NA | 46 - 65 | 57% female, 43% male | supplement tables 1 + 2 |
| 51 | 9/9/2020 | 10/9/2020 | 9/24/2020 | NA | Euroimmun anti-S | NA | 46 - 65 | 57% female, 43% male | supplement tables 1 + 2 |
| 51 | 10/15/2020 | 11/15/2020 | 10/30/2020 | NA | Euroimmun anti-S | NA | 46 - 65 | 57% female, 43% male | supplement tables 1 + 2 |
| 51 | 11/18/2020 | 12/15/2020 | 12/1/2020 | NA | Euroimmun anti-S | NA | 46 - 65 | 57% female, 43% male | supplement tables 1 + 2 |
| 51 | 10/14/2020 | 11/10/2020 | 10/27/2020 | NA | Euroimmun anti-S | NA | 46 - 65 | 57% female, 43% male | supplement tables 1 + 2 |
| 51 | 11/18/2020 | 12/15/2020 | 12/1/2020 | NA | Euroimmun anti-S | NA | 46 - 65 | 57% female, 43% male | supplement tables 1 + 2 |
| 51 | 1/25/2021 | 2/27/2021 | 2/10/2021 | NA | Euroimmun anti-S | NA | 46 - 65 | 57% female, 43% male | supplement tables 1 + 2 |
| 51 | 3/1/2021 | 3/31/2021 | 3/16/2021 | NA | Euroimmun anti-S | NA | 46 - 65 | 57% female, 43% male | supplement tables 1 + 2 |
| 51 | 4/7/2021 | 5/5/2021 | 4/21/2021 | NA | Euroimmun anti-S | NA | 46 - 65 | 57% female, 43% male | supplement tables 1 + 2 |

| | | | | | | | | | |
|---|---|---|---|---|---|---|---|---|---|
| 51 | 3/3/2021 | 8/17/2021 | 5/25/2021 | NA | Euroimmun anti-S | NA | 46 - 65 | 57% female, 43% male | supplement tables 1 + 2 |
| 51 | 5/12/2021 | 6/8/2021 | 5/25/2021 | NA | Euroimmun anti-S | NA | 46 - 65 | 57% female, 43% male | supplement tables 1 + 2 |
| 52 , 57 | 9/8/2020 | 9/26/2020 | 9/17/2020 | NA | Euroimmun anti-S | NA | NA | NA | |
| 53 | NA | NA | NA | 1 | Euroimmun anti-S | NA | NA | NA | |
| 53 | NA | NA | NA | 1 - 3 | Euroimmun anti-S | NA | NA | NA | |
| 53 | NA | NA | NA | 1 | University of Rio de Janeiro | NA | NA | NA | |
| 53 | NA | NA | NA | 1 - 3 | University of Rio de Janeiro | NA | NA | NA | |
| 54 | 4/6/2020 | NA | NA | 2 | Euroimmun anti-S | 2.9% hospitalized | 44 | 54% female , 46% male | figure 1 |
| 54 | NA | NA | NA | 4 | Euroimmun anti-S | 2.9% hospitalized | 44 | 54% female , 46% male | figure 1 |
| 54 | NA | NA | NA | 7 | Euroimmun anti-S | 2.9% hospitalized | 44 | 54% female , 46% male | figure 1 |
| 54 | NA | 12/17/2020 | NA | 9 | Euroimmun anti-S | 2.9% hospitalized | 44 | 54% female , 46% male | figure 1 |
| 57 | 6/23/2020 | 7/4/2020 | 6/28/2020 | NA | Euroimmun anti-S | NA | NA | NA | |
| 57 | 11/17/2020 | 12/5/2020 | 11/26/2020 | NA | Euroimmun anti-S | NA | NA | NA | |
| 58 | NA | NA | 11/11/2020 | 2 | Euroimmun anti-S | NA | 35 - 49 | 47% female , 53% male | table 1 |
| 59 | 6/20/2020 | 7/13/2020 | 7/1/2020 | NA | Fortress Diagnostics | NA | NA | NA | |
| 60 | 7/31/2020 | 8/13/2020 | 8/6/2020 | NA | Fortress Diagnostics | NA | NA | NA | |
| 60 | 9/15/2020 | 9/28/2020 | 9/21/2020 | NA | Fortress Diagnostics | NA | NA | NA | |
| 61 | 10/27/2020 | 11/10/2020 | 11/3/2020 | NA | Fortress Diagnostics | NA | NA | NA | |
| 62 | 4/27/2020 | 5/29/2020 | 5/13/2020 | NA | Hangzhou Clongene | NA | NA | NA | |
| 63 | NA | NA | October 2020 | 8 | THL Finland, in-house FMIA | 13% hospitalized | 49 | 57% female , 43% male | tables 1 and 2 |

| | | | | | | | | | |
|---|---|---|---|---|---|---|---|---|---|
| 63 | NA | NA | May 2021 | 13 | THL Finland, in-house FMIA | 13% hospitalized | 49 | 57% female , 43% male | tables 1 and 2 |
| 63 | NA | NA | October 2020 | 8 | THL Finland, in-house neutralisation assay | 13% hospitalized | 49 | 57% female , 43% male | tables 1 and 2 |
| 63 | NA | NA | May 2021 | 13 | THL Finland, in-house neutralisation assay | 13% hospitalized | 49 | 57% female , 43% male | tables 1 and 2 |
| 64 | NA | NA | NA | 4 | THL Finland, in-house neutralisation assay | NA | NA | NA | |
| 65 | early November 2020 | December 2020 | NA | >6 | THL Finland, in-house neutralisation assay | NA | NA | NA | |
| 66 | 11/4/2020 | 1/22/2021 | 12/13/2020 | NA | Gauteng, in-house Luminex | NA | NA | NA | |
| 67 | March 2021 | April 2021 | | NA | Tunis, in-house | NA | NA | NA | |
| 68 | 5/4/2020 | 5/13/2020 | 5/8/2020 | NA | Livzon lateral flow | NA | NA | NA | |
| 69 | 10/2/2020 | 10/11/2020 | 10/6/2020 | 1 | Livzon lateral flow | NA | 49 [34 - 59] | 61% female, 39% male | table 2 |
| 69 | 10/2/2020 | 10/11/2020 | 10/6/2020 | 2 | Livzon lateral flow | NA | 49 [34 - 59] | 61% female, 39% male | table 2 |
| 69 | 10/2/2020 | 10/11/2020 | 10/6/2020 | 4 | Livzon lateral flow | NA | 49 [34 - 59] | 61% female, 39% male | table 2 |
| 69 | 10/2/2020 | 10/11/2020 | 10/6/2020 | 6 | Livzon lateral flow | NA | 49 [34 - 59] | 61% female, 39% male | table 2 |
| 69 | 10/2/2020 | 10/11/2020 | 10/6/2020 | 7 | Livzon lateral flow | NA | 49 [34 - 59] | 61% female, 39% male | table 2 |
| 70 | 4/9/2021 | 4/25/2021 | 4/17/2021 | NA | Luminex xMAP (RBD portion) | NA | NA | NA | |
| 71 | 11/5/2020 | 12/31/2020 | 12/3/2020 | NA | Massachusetts General Hospital | NA | NA | NA | |
| 72 | October 2020 | March 2021 | | NA | Mount Sinai (New York) | ≤ 6% hospitalized | NA | NA | |
| 72 | October 2020 | March 2021 | | >6 | Mount Sinai (New York) | ≤ 27% hospitalized | NA | NA | |

| | | | | | | | | | |
|---|---|---|---|---|---|---|---|---|---|
| 73 | 5/18/2020 | 6/1/2020 | 5/25/2020 | NA | Orient Gene Biotech | NA | NA | 58% female , 42% male | tabla 2 on page 10 |
| 73 | 6/8/2020 | 6/22/2020 | 6/15/2020 | NA | Orient Gene Biotech | NA | NA | 58% female , 42% male | tabla 2 on page 10 |
| 74 | 11/16/2020 | 11/29/2020 | 11/22/2020 | NA | Orient Gene Biotech | NA | NA | 54% female , 46% male | tabla 2 on page 11 |
| 75 | October 2020 | February 2021 | December 2020 | NA | SenASTrIS test | NA | NA | NA | |
| 76 | 5/4/2020 | 6/27/2020 | 5/31/2020 | 2 - 3 | SenASTrIS test | 15% hospitalized | 49 | 56% female , 44% male | table 1 |
| 77 | 9/8/2020 | 10/14/2020 | 9/26/2020 | NA | Siemens Advia Centaur | NA | NA | NA | |
| 78 | 6/28/2020 | 7/9/2020 | 7/3/2020 | NA | Standard Q | NA | NA | NA | |
| 79 | NA | NA | NA | 2 - 3 | Sinai Health (Toronto) | ~26% hospitalized | 58 | 48% female , 52% male | table 1, figure 1 legend |
| 80 | 12/7/2020 | 1/15/2021 | 12/26/2020 | NA | Techno Genetics | NA | NA | NA | |
| 81 | 6/10/2020 | 7/29/2020 | 7/4/2020 | NA | Vitros Ortho (not total antibodies) | NA | NA | NA | |
| 82 | 1/11/2021 | 1/22/2021 | 1/16/2021 | NA | Vitros Ortho (not total antibodies) | NA | NA | NA | |
| 83 | 9/24/2021 | 10/14/2021 | 10/4/2021 | NA | Vitros Ortho (not total antibodies) | NA | NA | NA | |
| 84 | 10/15/2020 | 11/16/2020 | 10/31/2020 | 4 | Vitros Ortho (not total antibodies) | NA | NA | NA | |
| 84 | 4/18/2020 | 4/19/2020 | 4/18/2020 | 1 | Wantai | NA | NA | NA | |
| 86 | 5/18/2020 | 12/16/2020 | 9/1/2020 | 2 | Wantai | NA | NA | NA | |
| 87 | 2/27/2021 | 4/20/2021 | 3/17/2021 | 4 | Wantai | NA | NA | NA | |
| 88 | 5/14/2021 | 7/11/2021 | 6/10/2021 | 5 | Wantai | NA | NA | NA | |

#### B) Supplementary methods, statistical modeling

Cross validation posterior sample generation

Because the use of our estimates to predict sensitivity for serosurveys in the literature may involve extrapolating our curves through time (e.g. if an assay has data up to a limited time range), we used a cross-validation procedure that shows model performance with extrapolation through time. We achieved this by grouping the validation data points so that validation involved extrapolation through time.

First we selected an assay to be validated. Then, we computed the average of the times from diagnosis to serological testing reported across the assay's data points. We next separated the assay's data points into those with times earlier, and those with times later than the average time (i.e. cohorts serologically tested closer to their diagnosis date, and further to their diagnosis, respectively). We then fitted the model to all the data, except the data points with earlier times. We used this model fit to generate predictions of the sensitivities of the excluded data points (i.e. estimate the sensitivity of a given assay, at a given time). We repeated the procedure fitting all the data except the "later" data points for this assay (we now included the "earlier" data points for this assay in the model fit). We repeated this procedure for all assays, thus generating predictions for all the data points included in the analysis.

For the generation of the posterior samples, say that we want to estimate the number of positive tests $x_{a,s,t}$ for a left out data point corresponding to assay $a$, in study $s$, at time $t$. Suppose that other data points of the same assay $a$ and of the same study $s$ as this data point were used in model fitting. Then, at each posterior draw of the model, we have samples of the mean intercept and slope across assays, $\mu$ and $\beta$, samples of the assay specific intercept and slope effects $u_a$ and $b_a$, and a sample of the study specific effect $u_s$. Then, we plug these parameter samples into equation (2) of the Methods in the main text, to obtain an assay and study specific sample of sensitivity at time $t$, $\theta_{a,s,t}$. Next, we use this sample of $\theta_{a,s,t}$ to proceed as explained in the main methods, sampling $x_{a,s,t}$ 5 times from binomial distribution in equation (1) with parameters $\theta_{a,s,t}$ and $N_{a,s,t}$. After repeating this procedure for all posterior samples of the parameters, we take the 2.5% and 97.5% quantiles of the $x_{a,s,t}$ samples as the 95% CrI of our prediction.

In the case of $x_{a,s,t}$ where no other sample from the same study was used in model fitting, we do not have posterior draws of the study-specific effect $u_s$ generated by the model. For these cases, at each draw, we sample a value of $u_s$ from the distribution $N(u_s|0, \sigma_{us})$, using the corresponding draw of $\sigma_{us}$, and then follow the procedure described above. Similarly, in the cases where $x_{a,s,t}$ is the only data point for an assay and there are no assay-specific effects $u_a$ and $b_a$ in the posterior sample, we sample these from $N(u_a|0, \sigma_{ua})$ and $N(b_a|0, \sigma_{ba})$ respectively. That is, for assays with only one data point, we only sample sensitivities from the overall distribution of sensitivities.

To obtain model predictions, we first sampled the sensitivity of a given data point from the fitted Bayesian model as described above. Then, for each posterior draw of sensitivity, we drew 5 samples of the number of positive tests using the sampled sensitivity and Equation 1 of the main text. This produced a posterior distribution of the number of positive tests for each data point.

We apply an analogous procedure to cross-validate the models that include effects of assay characteristics.

Estimation of times between diagnosis and serology

When studies did not report the median time between diagnosis and serological testing for their cohort, we estimated these times using reported case curves for the study's location. For this, we first took the cumulative number of reported cases *C* at the study's location at the date of serological sampling, and set the median date of COVID-19 diagnosis for the tested cohort as the date in which the cumulative cases were *C*/2 in this location. If the study's cohort is representative of the general population, the population's median diagnosis date should match that of the cohort. We then took the time elapsed between this median diagnosis date and the median serosurvey sampling date as the time from diagnosis to sampling for this data point. Case numbers were obtained from (90), or from data made available by public health authorities.

#### C) Assay-specific sensitivity profiles

With the models fitted to sensitivity data, we obtained the posterior probability of time-varying sensitivity across tests. **Figure S1** shows the estimated time-dependent sensitivity for each assay, the fitted data (each point in **Figure S1** corresponds to a row in **Table S2**), and we indicate which data points were within the 95% CrI of their cross-validated prediction. We show the sensitivity profiles predicted from the model without assay characteristics (black) and with assay characteristics (red). We also present the values estimated for time-varying sensitivity across 14 months for each assay in **Table S3** (i.e. the values corresponding to the red lines and their credible intervals).

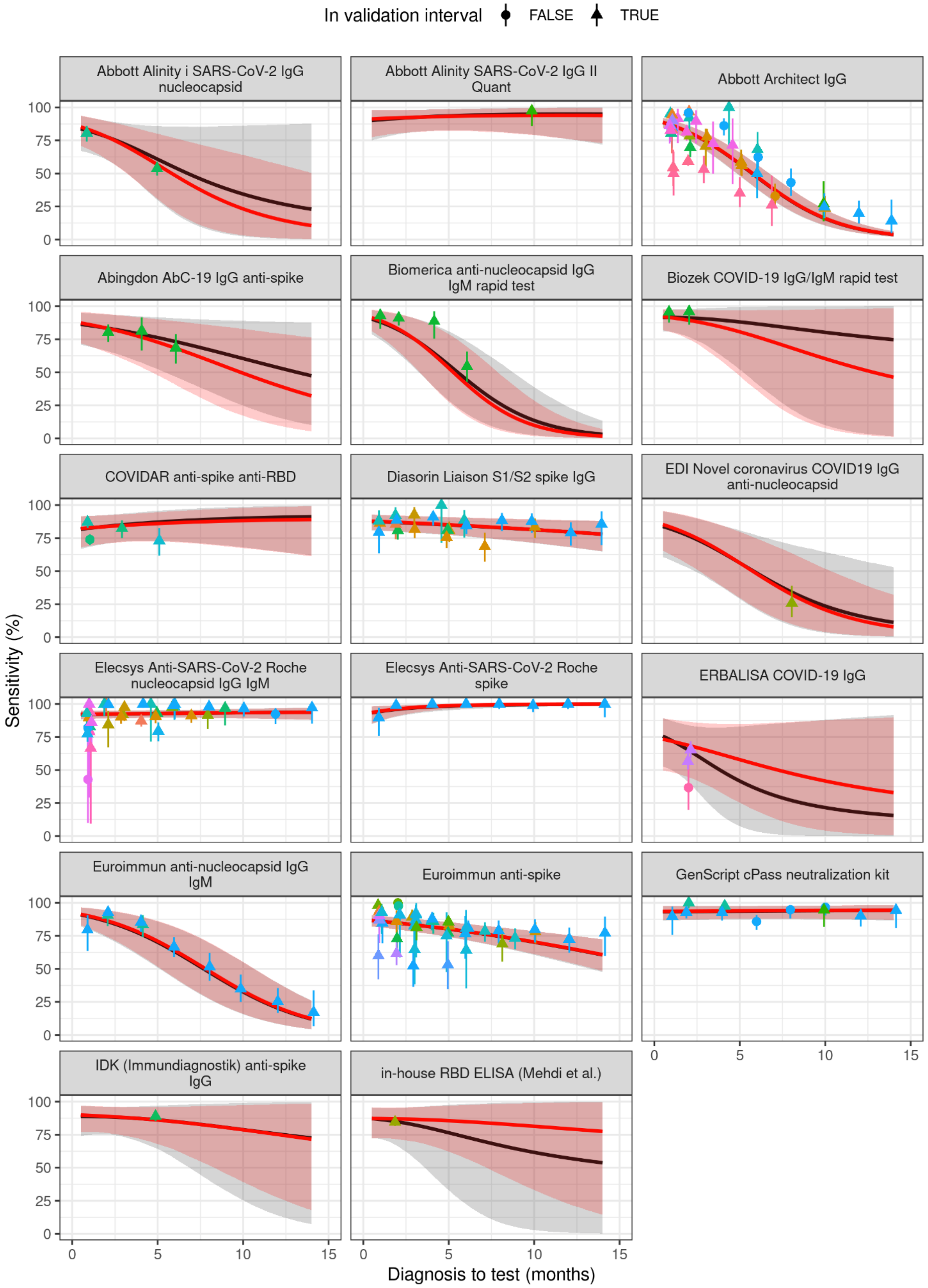
In validation interval
FALSE
TRUE
Abbott Alinity i SARS-CoV-2 IgG nucleocapsid
Abbott Alinity SARS-CoV-2 IgG II Quant
Abbott Architect IgG
Abingdon AbC-19 IgG anti-spike
Biomerica anti-nucleocapsid IgG IgM rapid test
Biozek COVID-19 IgG/IgM rapid test
COVIDAR anti-spike anti-RBD
Diasorin Liaison S1/S2 spike IgG
EDI Novel coronavirus COVID19 IgG anti-nucleocapsid
Elecsys Anti-SARS-CoV-2 Roche nucleocapsid IgG IgM
Elecsys Anti-SARS-CoV-2 Roche spike
ERBALISA COVID-19 IgG
Euroimmun anti-nucleocapsid IgG IgM
Euroimmun anti-spike
GenScript cPass neutralization kit
IDK (Immundiagnostik) anti-spike IgG
in-house RBD ELISA (Mehdi et al.)
Sensitivity (%)
Diagnosis to test (months)
0
25
50
75
100
0
5
10
15

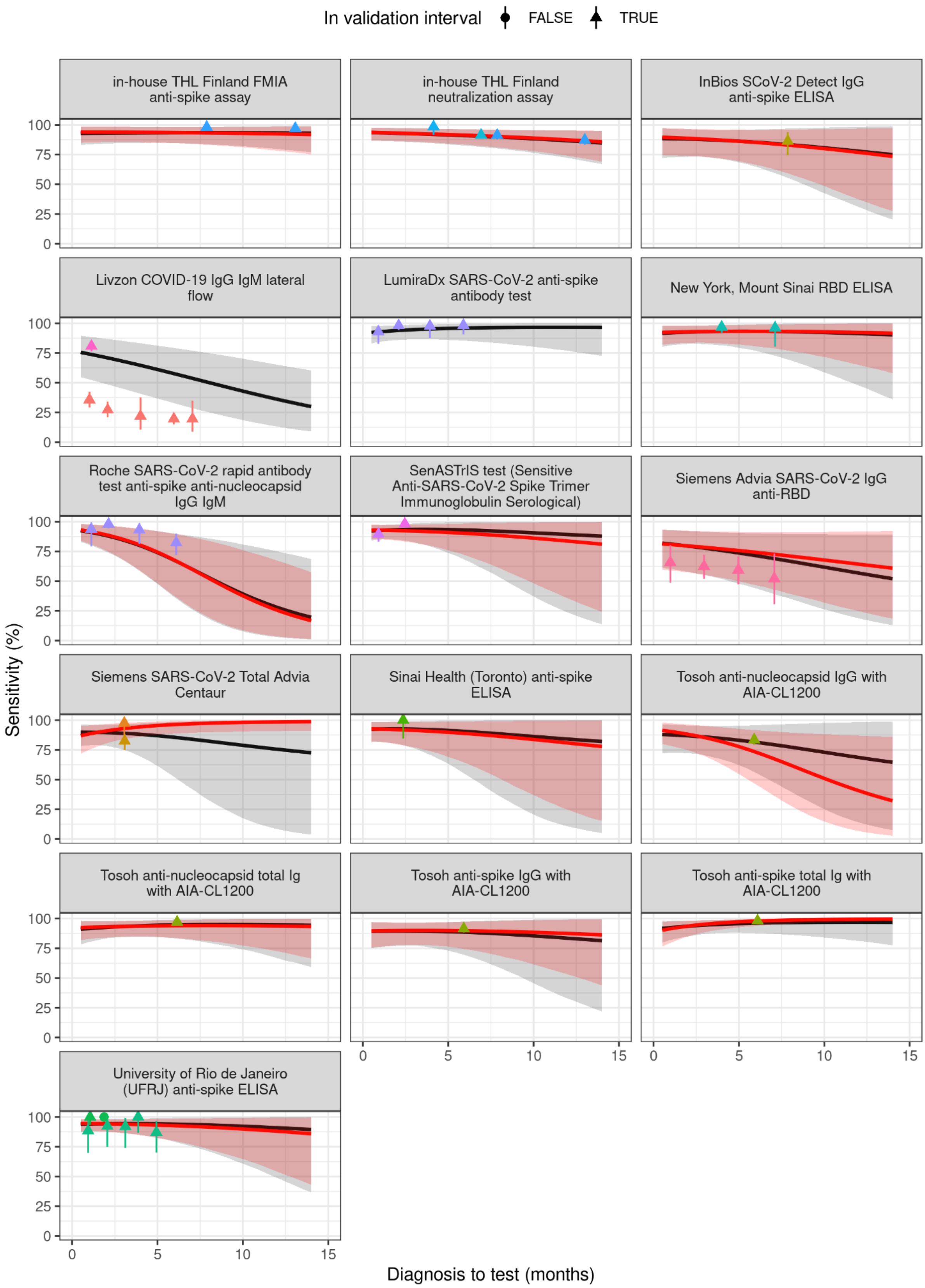
In validation interval
FALSE
TRUE
in-house THL Finland FMIA anti-spike assay
in-house THL Finland neutralization assay
InBios SCoV-2 Detect IgG anti-spike ELISA
Livzon COVID-19 IgG IgM lateral flow
LumiraDx SARS-CoV-2 anti-spike antibody test
New York, Mount Sinai RBD ELISA
Roche SARS-CoV-2 rapid antibody test anti-spike anti-nucleocapsid IgG IgM
SenASTrIS test (Sensitive Anti-SARS-CoV-2 Spike Trimer Immunoglobulin Serological)
Siemens Advia SARS-CoV-2 IgG anti-RBD
Siemens SARS-CoV-2 Total Advia Centaur
Sinai Health (Toronto) anti-spike ELISA
Tosoh anti-nucleocapsid IgG with AIA-CL1200
Tosoh anti-nucleocapsid total Ig with AIA-CL1200
Tosoh anti-spike IgG with AIA-CL1200
Tosoh anti-spike total Ig with AIA-CL1200
University of Rio de Janeiro (UFRJ) anti-spike ELISA
Sensitivity (%)
0
25
50
75
100
Diagnosis to test (months)
0
5
10
15

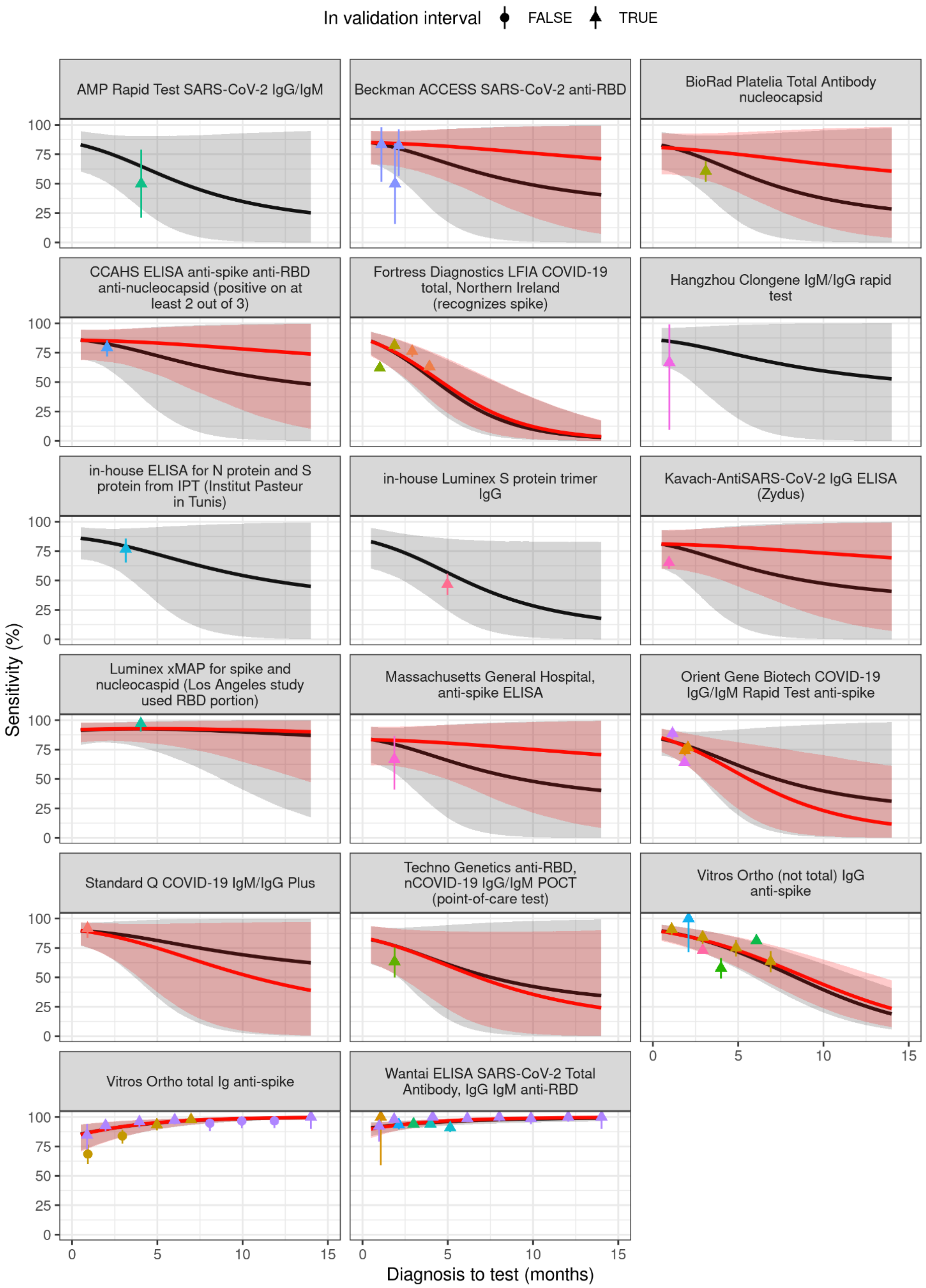

**Figure S1. Assay-specific sensitivity profiles**. Sensitivity across time estimated for all fitted assays. Each panel shows a different antibody assay. Points show the fitted data for each assay, with vertical bars showing the 95% CI of each point. Point shape indicates whether the data point was inside their prediction interval in cross-validation (triangles) or not (circles). Black lines show the mean estimated sensitivity for each assay using the model without assay characteristics. The gray shaded region indicates the 95% posterior interval for the sensitivity estimates. Red lines and shaded regions show the same for the full model (taking into account assay antigen and analytical technique). Assays lacking a red line were not included in the full model fit, due to the absence of assay characteristics data. The color of the data points indicates the study that reported the data point, for easier identification of data points belonging to the same study.

**Table S3. Time-varying sensitivity estimates for each assay.** In this table we show the estimated sensitivities and 95% CrI (for the average assay sensitivity, not counting for between-study variability) for each assay included in our analysis. For assays that had sufficient information to fit the full model (e.g. do not lack information about the targeted antigen), we report the sensitivities obtained from the full model. If we lacked information about some assay characteristic, we report the sensitivities estimated with a model fitted without that characteristic. Assays that were not fit in the full model are indicated by an asterisk (*) after their name.

| Assay | Month | | | | | | | | | | | | | |
|---|---|---|---|---|---|---|---|---|---|---|---|---|---|---|
| | 1 | 2 | 3 | 4 | 5 | 6 | 7 | 8 | 9 | 10 | 11 | 12 | 13 | 14 |
| Abbott Alinity i | 83 (68-92) | 78 (61-90) | 72 (52-87) | 64 (41-83) | 57 (31-80) | 49 (21-77) | 41 (14-73) | 34 (9-70) | 28 (5-67) | 23 (3-64) | 19 (2-61) | 16 (1-57) | 13 (1-54) | 11 (0-50) |
| Abbott Quant II | 92 (79-98) | 92 (80-98) | 93 (81-98) | 93 (82-98) | 93 (82-99) | 93 (82-99) | 94 (82-99) | 94 (81-99) | 94 (80-99) | 94 (79-99) | 94 (77-100) | 94 (76-100) | 94 (75-100) | 94 (73-100) |
| Abbott Architect | 87 (82-90) | 81 (76-86) | 75 (68-81) | 67 (58-74) | 57 (49-66) | 48 (39-57) | 38 (29-47) | 29 (22-38) | 22 (15-30) | 16 (11-23) | 11 (7-17) | 8 (5-12) | 6 (3-9) | 4 (2-6) |
| Abingdon AbC-19 | 86 (70-95) | 83 (65-94) | 80 (60-93) | 77 (54-92) | 73 (48-90) | 68 (41-89) | 64 (34-87) | 59 (28-86) | 54 (22-84) | 49 (17-83) | 45 (13-81) | 40 (10-79) | 36 (7-78) | 32 (5-76) |
| Beckman ACCESS | 85 (66-95) | 84 (65-95) | 83 (62-95) | 83 (57-96) | 82 (52-96) | 81 (46-97) | 80 (39-97) | 78 (33-98) | 77 (27-98) | 76 (22-99) | 75 (17-99) | 73 (13-99) | 72 (10-99) | 71 (7-99) |
| Biomerica | 89 (77-97) | 83 (67-94) | 75 (54-91) | 65 (40-86) | 53 (28-79) | 41 (18-70) | 31 (10-60) | 22 (6-48) | 15 (3-38) | 10 (2-29) | 6 (1-21) | 4 (1-15) | 3 (0-10) | 2 (0-7) |
| BioRad Platelia | 80 (58-93) | 79 (57-92) | 78 (53-93) | 77 (49-93) | 75 (43-94) | 73 (36-94) | 72 (29-95) | 70 (23-96) | 68 (18-96) | 67 (14-97) | 65 (10-97) | 63 (7-98) | 62 (5-98) | 61 (4-98) |
| Biozek | 91 (80-98) | 89 (75-97) | 87 (67-97) | 84 (58-97) | 81 (47-97) | 77 (37-97) | 73 (26-97) | 69 (18-97) | 64 (12-97) | 60 (8-98) | 56 (5-98) | 53 (3-98) | 49 (2-98) | 46 (1-98) |

| | | | | | | | | | | | | | | |
|---|---|---|---|---|---|---|---|---|---|---|---|---|---|---|
| CCAHS ELISA | 85 (69-95) | 85 (67-95) | 85 (65-95) | 84 (61-96) | 83 (56-96) | 82 (50-97) | 81 (44-98) | 80 (38-98) | 79 (32-98) | 78 (26-99) | 77 (21-99) | 76 (17-99) | 75 (13-99) | 74 (10-99) |
| COVIDAR | 83 (69-92) | 84 (71-92) | 85 (72-93) | 85 (72-94) | 86 (72-95) | 87 (72-95) | 87 (71-96) | 88 (70-97) | 88 (69-97) | 88 (67-98) | 89 (66-98) | 89 (65-99) | 89 (63-99) | 89 (61-99) |
| DiaSorin | 88 (81-93) | 87 (80-92) | 86 (79-92) | 86 (79-91) | 85 (78-91) | 84 (77-91) | 84 (75-90) | 83 (74-90) | 82 (73-90) | 81 (71-89) | 81 (70-89) | 80 (68-89) | 79 (67-88) | 78 (65-88) |
| Epitope Diagnostics, Inc. | 83 (62-94) | 78 (55-92) | 72 (47-88) | 64 (38-84) | 56 (30-80) | 48 (22-75) | 40 (15-69) | 33 (10-64) | 26 (6-58) | 21 (4-52) | 16 (2-46) | 13 (1-41) | 10 (1-36) | 8 (1-32) |
| Roche Elecsys N | 92 (89-95) | 92 (89-95) | 93 (89-95) | 93 (89-95) | 93 (90-95) | 93 (90-95) | 93 (90-96) | 93 (89-96) | 93 (89-96) | 93 (89-96) | 93 (89-97) | 94 (89-97) | 94 (88-97) | 94 (88-97) |
| Roche Elecsys S | 95 (87-98) | 96 (91-99) | 97 (93-99) | 98 (95-100) | 99 (96-100) | 99 (97-100) | 99 (98-100) | 99 (98-100) | 100 (99-100) | 100 (99-100) | 100 (99-100) | 100 (99-100) | 100 (99-100) | 100 (99-100) |
| ErbaLisa | 72 (48-88) | 69 (44-87) | 65 (39-85) | 62 (32-85) | 58 (25-85) | 54 (19-85) | 51 (14-86) | 48 (9-86) | 44 (6-87) | 42 (4-88) | 39 (3-88) | 37 (2-89) | 35 (1-89) | 33 (1-90) |
| Euroimmun N | 90 (80-96) | 87 (75-94) | 82 (68-92) | 77 (60-90) | 71 (52-86) | 64 (43-82) | 56 (35-77) | 48 (28-70) | 41 (21-63) | 33 (16-55) | 27 (12-47) | 21 (9-39) | 16 (6-32) | 12 (4-26) |
| Euroimmun S1 | 86 (81-90) | 85 (80-89) | 83 (78-88) | 82 (76-87) | 80 (74-85) | 78 (72-84) | 77 (70-83) | 75 (67-81) | 73 (64-80) | 70 (61-79) | 68 (58-77) | 66 (55-76) | 63 (52-74) | 61 (48-72) |
| Fortress Diagnostics | 82 (68-91) | 75 (59-87) | 66 (47-82) | 56 (35-76) | 46 (24-70) | 37 (15-63) | 29 (9-56) | 22 (5-50) | 16 (3-43) | 12 (2-37) | 9 (1-31) | 7 (0-26) | 5 (0-22) | 4 (0-18) |
| GenScript cPass | 94 (87-98) | 94 (87-98) | 94 (88-98) | 94 (88-98) | 94 (88-98) | 94 (88-98) | 94 (88-98) | 94 (88-98) | 94 (88-98) | 94 (88-98) | 94 (88-98) | 94 (88-98) | 94 (87-98) | 94 (87-98) |
| Immundiagnostik | 90 (77-97) | 89 (76-96) | 88 (74-96) | 87 (72-96) | 86 (68-96) | 85 (64-96) | 83 (58-96) | 82 (52-97) | 80 (45-97) | 79 (39-97) | 77 (33-98) | 75 (28-98) | 73 (22-98) | 72 (18-98) |

| | | | | | | | | | | | | | | |
|---|---|---|---|---|---|---|---|---|---|---|---|---|---|---|
| Mehdi, in-house | 87 (72-95) | 87 (72-96) | 87 (70-96) | 86 (67-96) | 86 (62-97) | 85 (57-97) | 84 (51-98) | 83 (45-98) | 82 (39-99) | 81 (33-99) | 80 (28-99) | 79 (23-99) | 78 (18-99) | 78 (14-100) |
| THL Finland FMIA | 94 (85-99) | 94 (86-99) | 94 (86-98) | 94 (85-98) | 94 (85-98) | 94 (85-98) | 93 (84-98) | 93 (83-98) | 93 (82-98) | 93 (81-98) | 93 (80-98) | 92 (78-98) | 92 (77-99) | 92 (75-99) |
| THL Finland neutralizat ion | 93 (87-98) | 93 (86-97) | 93 (86-97) | 92 (85-97) | 92 (84-97) | 91 (83-96) | 91 (82-96) | 90 (81-96) | 90 (79-96) | 89 (78-96) | 88 (76-95) | 87 (74-95) | 87 (72-95) | 86 (69-95) |
| InBios SCoV-2 Detect | 89 (74-97) | 89 (74-97) | 88 (73-96) | 87 (72-96) | 86 (69-96) | 85 (66-96) | 84 (63-96) | 83 (59-96) | 81 (54-96) | 80 (49-96) | 78 (43-97) | 77 (37-97) | 75 (32-97) | 74 (28-97) |
| Zydus Kavach ELISA IgG | 81 (60-93) | 80 (58-93) | 80 (55-94) | 79 (51-95) | 78 (45-96) | 77 (39-96) | 76 (34-97) | 75 (28-98) | 74 (23-98) | 73 (19-98) | 72 (15-99) | 71 (12-99) | 70 (9-99) | 69 (7-99) |
| Luminex xMAP, RBD portion | 92 (82-98) | 92 (82-98) | 93 (82-98) | 93 (81-98) | 93 (80-99) | 93 (78-99) | 93 (75-99) | 92 (72-99) | 92 (69-99) | 92 (65-100) | 91 (60-100) | 91 (56-100) | 91 (52-100) | 90 (47-100) |
| Massachu setts General Hospital | 83 (62-94) | 83 (61-94) | 82 (58-95) | 81 (54-95) | 80 (50-96) | 79 (44-97) | 78 (38-97) | 77 (31-98) | 76 (26-98) | 75 (21-98) | 74 (17-99) | 73 (14-99) | 72 (11-99) | 71 (8-99) |
| Mount Sinai (New York) | 93 (83-98) | 93 (83-98) | 93 (84-98) | 93 (83-98) | 93 (82-99) | 93 (81-99) | 93 (79-99) | 93 (77-99) | 93 (75-99) | 93 (72-99) | 93 (68-100) | 92 (65-100) | 92 (62-100) | 92 (58-100) |
| Orient Gene | 83 (70-91) | 77 (61-89) | 70 (49-86) | 63 (36-83) | 55 (24-81) | 47 (15-79) | 40 (9-77) | 33 (5-75) | 28 (3-72) | 23 (1-70) | 19 (1-68) | 16 (0-65) | 14 (0-63) | 12 (0-61) |

| | | | | | | | | | | | | | | |
|---|---|---|---|---|---|---|---|---|---|---|---|---|---|---|
| Biotech | | | | | | | | | | | | | | |
| Roche, rapid test, S + N | 92 (82-98) | 89 (76-97) | 85 (68-96) | 80 (58-94) | 74 (48-92) | 68 (37-90) | 60 (28-87) | 52 (20-84) | 45 (13-80) | 38 (9-76) | 31 (6-72) | 26 (3-67) | 21 (2-62) | 17 (1-58) |
| SenASTrIS test | 93 (85-97) | 93 (84-97) | 92 (82-98) | 92 (80-98) | 91 (76-98) | 90 (72-98) | 89 (67-99) | 88 (61-99) | 87 (55-99) | 86 (48-99) | 85 (42-99) | 84 (36-100) | 82 (30-100) | 81 (24-100) |
| Siemens Advia SARS-CoV-2 IgG anti-RBD | 81 (59-93) | 80 (58-92) | 78 (55-92) | 77 (53-92) | 76 (50-91) | 74 (47-91) | 73 (43-91) | 71 (39-91) | 69 (36-91) | 68 (32-91) | 66 (28-92) | 64 (25-92) | 63 (22-92) | 61 (19-92) |
| Siemens Advia Centaur, Total | 88 (75-96) | 91 (80-97) | 93 (84-98) | 94 (86-98) | 95 (88-99) | 96 (89-99) | 97 (89-100) | 97 (90-100) | 98 (90-100) | 98 (90-100) | 98 (91-100) | 98 (91-100) | 99 (91-100) | 99 (91-100) |
| Sinai Health (Toronto) ELISA, S | 92 (82-98) | 92 (80-98) | 91 (77-98) | 91 (74-99) | 90 (69-99) | 89 (64-99) | 88 (58-99) | 86 (51-99) | 85 (44-99) | 84 (37-99) | 82 (30-99) | 81 (25-100) | 79 (20-100) | 78 (15-100) |
| SD Biosensor, Standard Q, Plus | 89 (75-96) | 86 (69-96) | 83 (60-96) | 79 (49-96) | 75 (37-96) | 70 (27-96) | 66 (18-96) | 61 (12-96) | 57 (8-96) | 52 (5-97) | 49 (3-97) | 45 (2-97) | 42 (1-97) | 39 (1-97) |
| Techno Genetics, anti-RBD POCT | 81 (59-93) | 76 (52-91) | 71 (43-90) | 66 (32-90) | 60 (23-89) | 54 (15-89) | 49 (10-89) | 44 (6-89) | 40 (3-89) | 36 (2-89) | 32 (1-89) | 29 (1-89) | 26 (0-90) | 24 (0-90) |
| Tosoh N indirect | 90 (78-97) | 88 (75-96) | 85 (70-95) | 82 (63-94) | 78 (55-93) | 73 (46-92) | 68 (37-90) | 62 (28-90) | 57 (21-89) | 51 (15-88) | 46 (10-88) | 41 (7-87) | 36 (4-86) | 32 (3-86) |

| | | | | | | | | | | | | | | |
|---|---|---|---|---|---|---|---|---|---|---|---|---|---|---|
| Tosoh N sandwich | 93 (83-98) | 93 (84-98) | 93 (85-98) | 94 (85-98) | 94 (85-98) | 94 (84-99) | 94 (83-99) | 94 (81-99) | 94 (79-99) | 94 (77-99) | 94 (75-100) | 94 (73-100) | 94 (70-100) | 93 (66-100) |
| Tosoh S indirect | 90 (76-97) | 90 (78-97) | 90 (78-97) | 90 (77-97) | 90 (76-97) | 90 (74-97) | 90 (72-98) | 89 (69-98) | 89 (66-98) | 88 (62-99) | 88 (57-99) | 87 (53-99) | 87 (49-99) | 86 (44-99) |
| Tosoh S sandwich | 92 (80-98) | 94 (85-98) | 95 (89-99) | 96 (91-99) | 97 (93-99) | 98 (94-100) | 98 (95-100) | 99 (96-100) | 99 (96-100) | 99 (97-100) | 99 (97-100) | 99 (97-100) | 100 (98-100) | 100 (98-100) |
| UFRJ (University of Rio de Janeiro) | 94 (88-98) | 94 (88-98) | 94 (86-98) | 94 (85-98) | 93 (83-98) | 93 (80-98) | 92 (77-99) | 91 (73-99) | 91 (69-99) | 90 (65-99) | 89 (60-99) | 88 (54-99) | 87 (49-99) | 86 (43-100) |
| Vitros Ortho, S CoV2 IgG | 88 (79-94) | 85 (75-92) | 82 (70-90) | 77 (65-87) | 73 (58-84) | 68 (51-81) | 62 (44-77) | 56 (37-74) | 50 (30-69) | 44 (24-65) | 38 (18-61) | 33 (14-56) | 28 (11-52) | 24 (8-47) |
| Vitros Ortho, RBD CoV2T total | 87 (73-94) | 90 (79-96) | 92 (83-97) | 94 (87-98) | 95 (89-98) | 96 (92-99) | 97 (93-99) | 98 (95-99) | 98 (96-99) | 99 (97-100) | 99 (97-100) | 99 (98-100) | 99 (98-100) | 100 (99-100) |
| Wantai ELISA | 91 (84-95) | 93 (88-96) | 95 (91-97) | 96 (93-98) | 97 (94-99) | 97 (95-99) | 98 (96-99) | 98 (96-100) | 99 (97-100) | 99 (97-100) | 99 (98-100) | 99 (98-100) | 99 (98-100) | 100 (98-100) |
| AMP, rapid test (*) | 82 (60-94) | 74 (49-91) | 66 (36-88) | 56 (24-85) | 46 (14-82) | 38 (7-79) | 30 (4-76) | 24 (2-74) | 19 (1-71) | 16 (0-68) | 13 (0-66) | 11 (0-63) | 9 (0-60) | 7 (0-58) |
| Clungene (*) | 81 (55-95) | 74 (42-94) | 66 (27-93) | 57 (15-92) | 49 (8-91) | 41 (4-91) | 35 (2-90) | 30 (1-90) | 26 (0-90) | 22 (0-90) | 19 (0-90) | 17 (0-89) | 15 (0-89) | 14 (0-89) |
| Livzon (*) | 74 (52-89) | 71 (48-87) | 67 (43-85) | 63 (39-83) | 60 (35-81) | 56 (31-78) | 52 (27-76) | 48 (23-73) | 44 (20-70) | 40 (17-67) | 37 (14-65) | 33 (12-62) | 30 (10-59) | 27 (8-56) |

| | | | | | | | | | | | | | | |
|---|---|---|---|---|---|---|---|---|---|---|---|---|---|---|
| Tunis, in-house (*) | 85 (67-94) | 85 (67-95) | 84 (65-95) | 84 (59-96) | 83 (52-97) | 82 (44-98) | 81 (36-98) | 80 (28-99) | 79 (22-99) | 78 (17-99) | 77 (12-100) | 76 (9-100) | 75 (6-100) | 74 (4-100) |
| Gauteng, in-house Luminex (*) | 79 (52-92) | 76 (51-91) | 74 (48-90) | 71 (43-90) | 67 (35-90) | 64 (28-91) | 60 (20-91) | 57 (14-92) | 54 (9-93) | 51 (6-94) | 49 (4-95) | 46 (3-95) | 44 (2-96) | 42 (1-96) |
| LumiraDx (*) | 93 (84-98) | 94 (86-98) | 95 (87-99) | 96 (88-99) | 96 (88-99) | 97 (88-100) | 97 (88-100) | 97 (88-100) | 98 (88-100) | 98 (87-100) | 98 (87-100) | 98 (86-100) | 98 (86-100) | 98 (85-100) |

#### D) Assay sensitivity increase can continue across months

One somewhat surprising result of the analyses in the main text is that several assays show positive slopes that can be interpreted as an increase of sensitivity throughout the months. This is surprising because while antibody levels rise in the early stages after infection, they tend to stabilize or drop afterwards (82,83). Although previous work has also reported increased sensitivity across the early months after infection for some assays (9,10,84), we wondered whether it is possible that after an initial increase in sensitivity for these assays, then sensitivity stabilizes or drops. We tested this possibility by fitting a new model with two slopes, an initial slope up to 3 months after infection, and a later slope afterwards. First, we filtered out assays that did not have sensitivity estimates both before and after the cutoff point. Then, out of the remaining assays, we fitted this model to the 7 assays that target RBD, and that do not use the LFA technique, since they show the most positive group slopes in **Figure 2** in the main text.

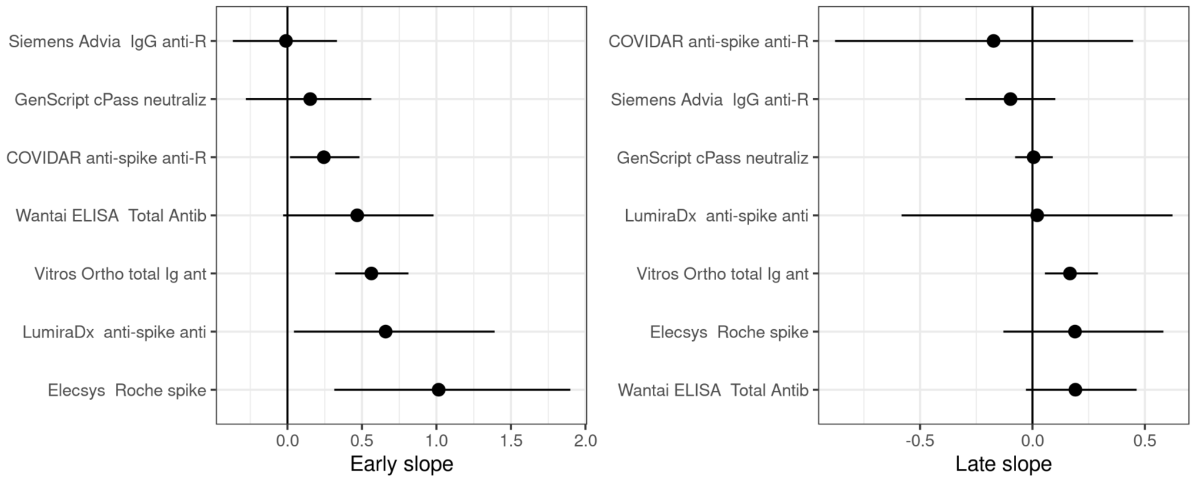

**Figure S2. Values of early and late slopes.** Values of the slope between 0 and 3 months (left panel) and the slope from 3 months onwards (right panel) for non-LFA assays that target RBD. Points show the mean of the posterior distribution of the parameters, and horizontal bars show the 95% credible interval (CrI).

In the model with two slopes, we obtained an average early slope (from 0 to 3 months) of 0.44 (CrI95%: 0.08; 0.87), and an average late slope (3 months onwards) of 0.04 (CrI95%: -0.33; 0.39). The later slope was smaller than the earlier slope in 93.6% of the posterior samples. This supports the idea that the increase in sensitivity mostly occurs during the early stages post infection. However, some assays seem to maintain a positive slope even after 3 months post infection (**Figure S1**). For example, for the assays *Elecsys Anti-SARS-CoV-2 Roche spike*, *Vitros*

*Ortho total Ig anti-spike*, and *Wantai ELISA SARS-CoV-2 Total Antibody anti-RBD* assays, the assay-specific later slopes was larger than 0 in 86.4%, 99.8%, and 95.2% of the posterior samples. Thus, our analysis shows that some assays can show an increase in sensitivity even several months after initial infection, although it is expected that sensitivity drops eventually for these assays, given enough time after infection. Finally, we note that although one could think that reinfections may drive this increase in sensitivity, we find this possibility to be unlikely. On the one hand, reinfections are unlikely to be common in the first 3 months after infection in seropositive individuals (91), where we do observe a positive slope for the Quantitative-Direct RBD-targeting assays. Then, many of the studies were performed during 2020, when reinfections were still relatively uncommon.

#### E) Robustness to model architecture

To examine the robustness of the model presented in the main text, considering all relevant assay characteristics simultaneously, we analyze here each assay characteristic separately.

We first fitted the model containing only the terms for assay antigen. That is, the model had a different slope for each antigen. The posterior distributions of these parameters are shown in **Figure S3A**, and the sensitivity profiles are shown in **Figure S3B**. We find that N<S in 83.4% of the posterior samples (as suggested by **Figure S3A**). The mean slope of RBD had a positive value of 0.09 [-0.07; 0.23] with RBD>0 in 84.3% of the posterior samples. Also, RBD had the slowest seroreversion, with RBD>S in 98.2% of the posterior samples, RBD>N in 99.8% of the samples. These results are in agreement with the results reported in the main text, with the difference of showing a more significant difference between RBD and S targeting assays.

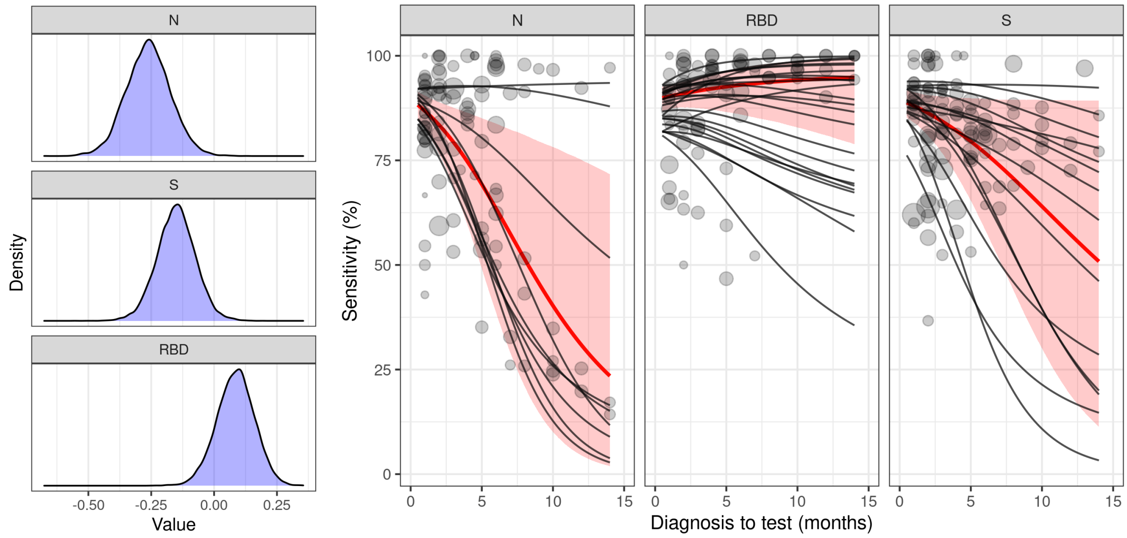

**Figure S3. Effect of antigen on sensitivity.** Estimated seroreversion segregated by the antigen used in the antibody assay. **A)** Posterior distribution of the effects of each antigen on the sensitivity slope. **B)** The sensitivity profile across time for each kind of assay is shown in a different panel. Panel titles indicate what slope parameters are added together to obtain the slope of each type of assay. The red lines show the mean sensitivity for each kind of assay across time, and the shaded regions show the 95% CrI. Note that the 95% CrI refer to the uncertainty over the mean sensitivity, and they do not account for the variability between assays. Gray lines show the fits for individual assays. Gray dots show the data, with dot size proportional to the square root of the number of samples.

Next, we looked at the effect of assay analytical technique. In particular, we fitted a different slope to each different type of analytic technique: LFA, Quantiative-Indirect, Quantitative-Direct and Competitive. In **Figure S4A** we see that the slopes for the different types of assays. The LFA slope had a value of -0.35 [-0.53; -0.19], the Quantitative-Indirect had a slope value of -0.12 [-0.22; -0.01], the Quantititative-Direct had a slope value of 0.20 [0.02; 0.38], and the Quantitative-Competitive had a slope value of -0.02 [-0.29; 0.25]. In **Figure S4B** we see the sensitivity across time profiles for these two kinds of assays. Similar to the results reported in the main text, we find that LFA assays have on average much faster resoreversion that quantitative assays. Also, within the quantitative assays, those using the direct binding design have more positive slopes, while there is considerable uncertainty in the model around the slope of assays using competitive binding (which in this case are neutralization assays).

We note that in further analysis (not shown), we did not find a clear difference between assays using ELISA, CLIA and CMIA types of detection method within the Quantitative assays.

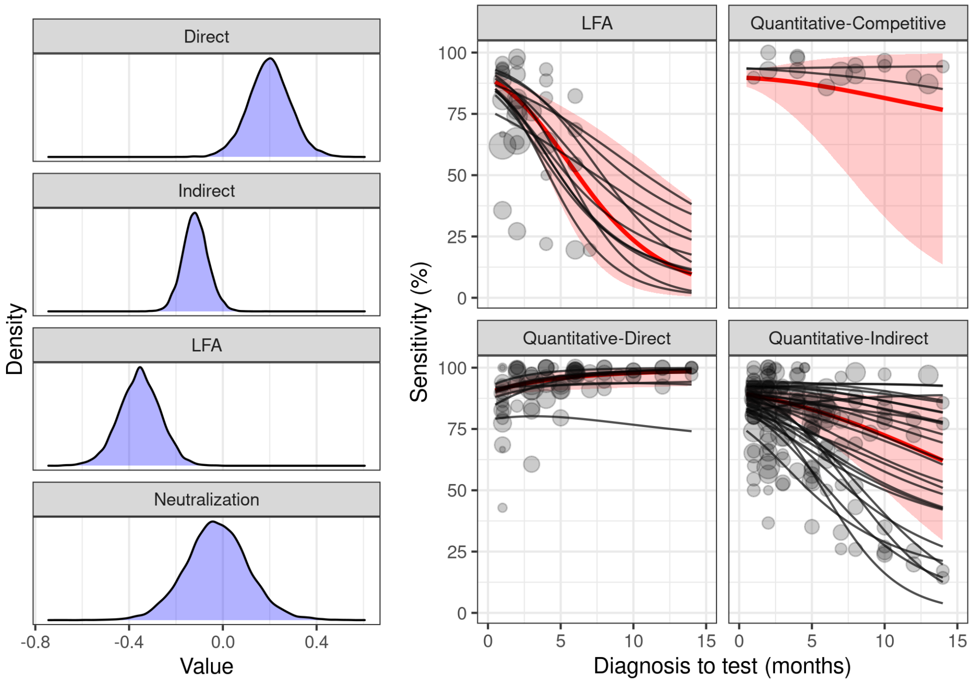

**Figure S4. Effect of analytic technique on sensitivity. A)** Posterior distribution of the mean slopes for each analytic technique. **B)** Sensitivity across time for each analytic technique. Same conventions as **Figure S3B**.

#### F) Robustness to time estimation

In our main analysis, we estimated the time between diagnosis and testing for samples where this number was unknown (see Methods). To test the robustness of the results to this procedure, we repeated the main analysis including only samples where the time between diagnosis and serology testing was known.

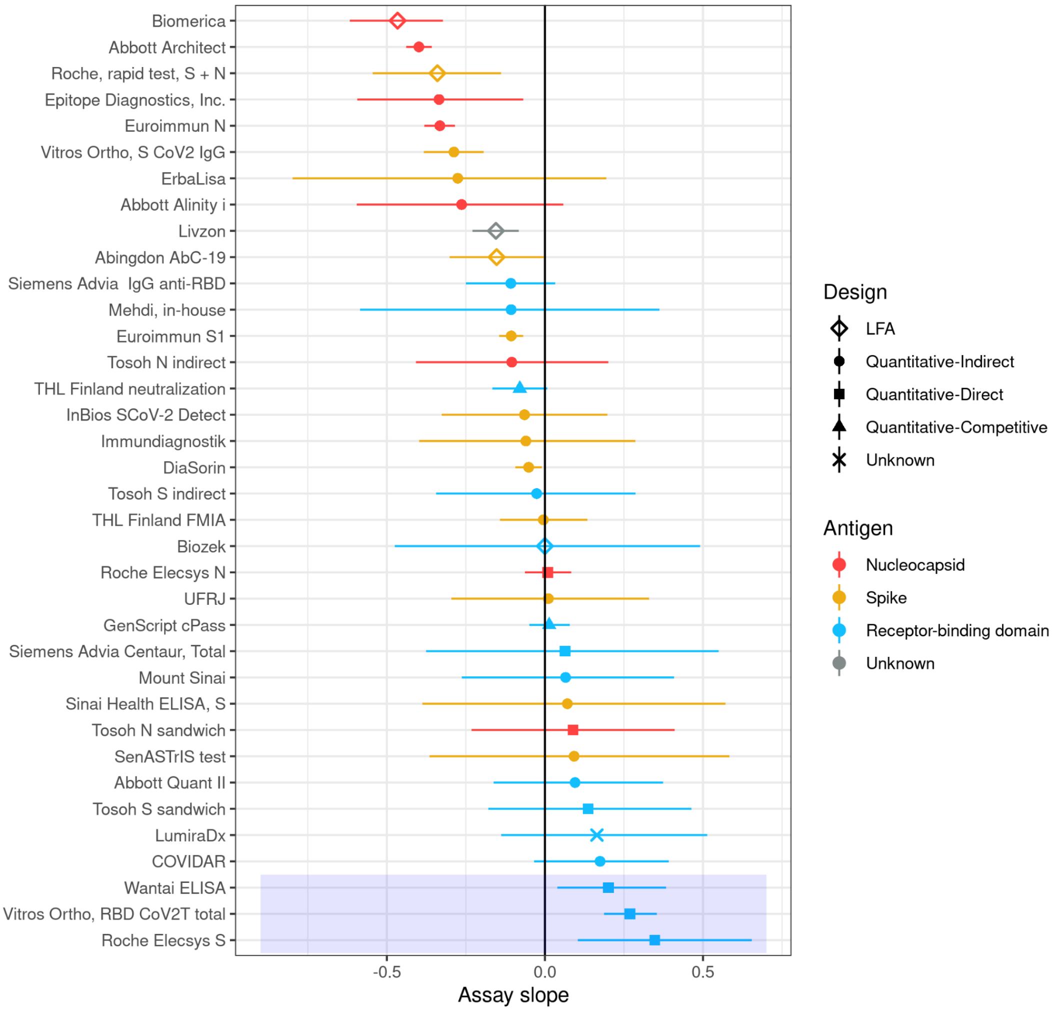

**Figure S5. Assay slopes estimated without assay characteristics, for samples with known times.** Same as **Figure 2** in the main text, but excluding the samples where the times between diagnosis and serology testing were estimated.

In **Figure S5** we see the slopes of the individual assays, analogous to **Figure 2** in the main text, and in **Figure S6** we see the average slopes for the different types of assays, analogous to **Figure 3** in the main text. We see that fewer assays are included in this analysis, because some assays only had data with unknown diagnosis to testing times. However, we see that the overall trends reported in the main text with the full dataset are the same as the ones obtained here with only known diagnosis to testing times. Estimated model parameters and statistical analyses are also in line with the main results (results not shown).

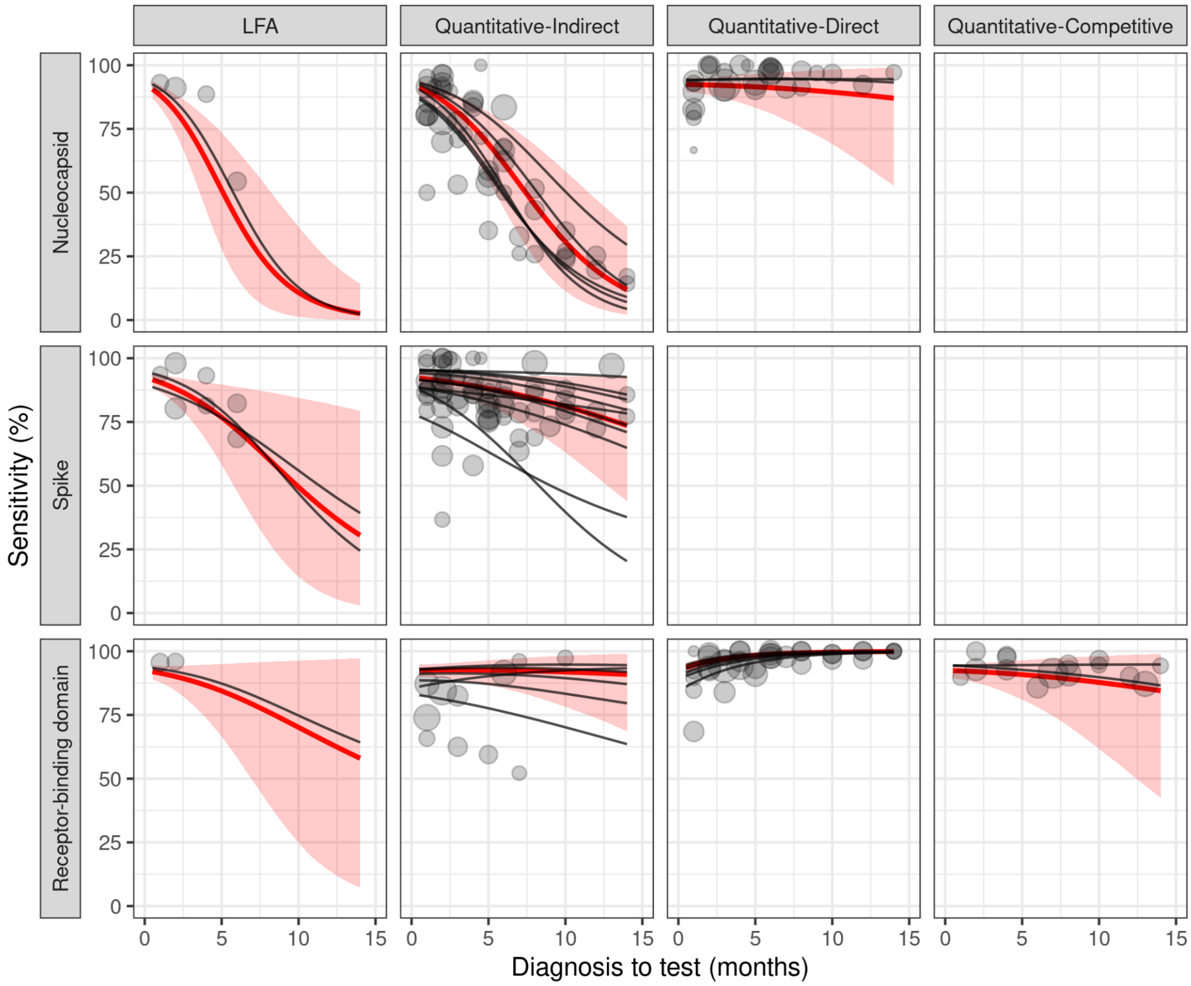

**Figure S6. Sensitivity profiles for different assay characteristics, for samples with known times.** Same as **Figure 3** in the main text, but excluding the samples where the times between diagnosis and serology testing were estimated.

#### G) Effect of assay characteristics on specificity

We fitted a hierarchical Bayesian model to specificity data reported for the assays included in our main analysis. Because assay specificity is not expected to have relevant dynamics (as opposed to sensitivity), we fitted only the point estimates of specificity reported for each assay by either the manufacturer, or independent studies. We fitted a model analogous to the model with assay characteristics discussed in the main text, with some modifications.

First, in the binomial distribution $P(x|\theta) \propto \theta^{x} \, (1-\theta)^{N-x}$ used to model the raw data, $N$ corresponds to the number of tested negative samples (e.g. pre-pandemic samples), and $x$ corresponds to the number of negative samples (i.e. the true negatives). In this context, parameter θ corresponds to assay specificity.

Then, we used the following formula to model average specificity by assay characteristics:

$$log\left(\frac{\theta_{a,s}}{1-\theta_{a,s}}\right) = \left(u_a + u_s + \beta_{LFA} L_a + \beta_{Direct} D_a + \beta_{Competitive} C_a + \beta_{Nucleocapsid} N_a + \beta_{Spike} S_a + \beta_{RBD} RBD_a\right)$$

In this model, we do not have a temporal component, all terms only affect the point value of specificity. We have an assay specific term $u_a$, a study specific term $u_s$, and the terms indicating the effects of different assay characteristics (e.g. $\beta_{LFA}$ the effect of being a qualitative, LFA assay), all analogous to the model described for sensitivity in the main text (e.g. we omit the Indirect assay term, making it the baseline specificity). Priors of the model parameters and the fitting procedure were set as for the model in the main text.

In **Figure S7** we see the raw specificity data, as well as the results obtained from the model fit. We obtained that LFA assays have on average lower specificity than Quantitative-Indirect assays ($\beta_{LFA}$<0 in 98.4% of the posterior samples), but we did not find a significant effect for the Quantitative-Direct design ($\beta_{Direct}$>0 in 85.0% of the posterior samples), the Quantitative-Competitive design ($\beta_{Competitive}$<0 in 86.6% of the posterior samples), or for the antigen targeted ($\beta_{Spike}$>$\beta_{Nucleocapsid}$ in 32.3% of the posterior samples, $\beta_{RBD}$>$\beta_{Nucleocapsid}$ in 67.3% of the posterior samples). Estimated sensitivities for different assay types are shown in **Table S4**.

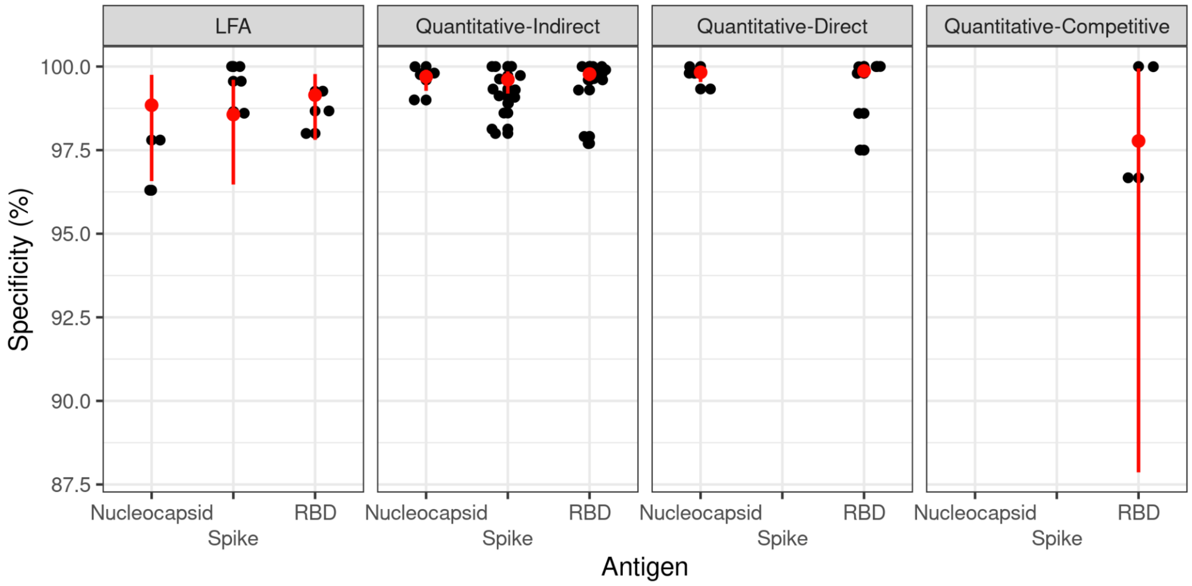

**Figure S7. Assay specificity for different assay characteristics.** Each black dot shows the specificity reported for a given assay, by a given study. Different panels indicate different test design, and the horizontal axis within each panel indicates the different antigens. Red dots and vertical lines indicate the mean sensitivity and 95% credible interval as obtained from the hierarchical Bayesian model fitted to the specificity data.

**Table S4.** Mean specificity and 95% CrI for the different types of assays, obtained from the fitted Bayesian model. Credible intervals do not account for variability between assays of a same type, or within studies. Values correspond to the red dots and lines in **Figure S7**.

| **Design** | **Antigen** | **Specificity % (95% CrI)** |
|---|---|---|
| LFA | N | 98.8 (96.6-99.7) |
| LFA | S | 98.6 (96.5-99.6) |
| LFA | RBD | 99.1 (97.8-99.8) |
| Quantitative-Indirect | N | 99.7 (99.3-99.9) |
| Quantitative-Indirect | S | 99.6 (99.1-99.9) |
| Quantitative-Indirect | RBD | 99.8 (99.6-99.9) |
| Quantitative-Direct | N | 99.8 (99.5-99.9) |
| Quantitative-Direct | RBD | 99.8 (99.7-100) |
| Quantitative-Competitive | RBD | 97.8 (87.9-99.9) |
